# Multiplexed biosensor for point-of-care COVID-19 monitoring: CRISPR-powered unamplified RNA diagnostics and protein-based therapeutic drug management

**DOI:** 10.1101/2022.03.30.22271928

**Authors:** Midori Johnston, H. Ceren Ates, Regina Glatz, Hasti Mohsenin, Rosanne Schmachtenberg, Nathalie Göppert, Daniela Huzly, Gerald A. Urban, Wilfried Weber, Can Dincer

## Abstract

In late 2019 SARS-CoV-2 rapidly spread to become a global pandemic, therefore, measures to attenuate chains of infection, such as high-throughput screenings and isolation of carriers were taken. Prerequisite for a reasonable and democratic implementation of such measures, however, is the availability of sufficient testing opportunities (beyond reverse transcription PCR, the current gold standard). We, therefore, propose an electrochemical, microfluidic multiplexed biosensor in combination with CRISPR/Cas-powered assays for point-of-care nucleic acid testing. In this study, we simultaneously screen for and identify SARS-CoV-2 infections (Omicron-variant) in clinical specimens (Sample-to-result time: ∼30 min), employing LbuCas13a, whilst bypassing reverse transcription as well as target amplification of the viral RNA, both of which are necessary for detection via PCR and multiple other methods. In addition, we demonstrate the feasibility of combining assays based on different classes of biomolecules, in this case protein-based antibiotic detection, on the same device. The programmability of the effector and multiplexing capacity (up to six analytes) of our platform, in combination with a miniaturized measurement setup, including a credit card sized near field communication (NFC) potentiostat and a microperistaltic pump, provide a promising on-site tool for identifying individuals infected with variants of concern and monitoring their disease progression alongside other potential biomarkers or medication clearance.

## Introduction

The severe acute respiratory syndrome coronavirus 2 (SARS-CoV-2) is a highly infectious member of the SARS-related Betacoronavirus (Sarbecovirus) family that probably emerged from a zoonotic reservoir in December of 2019^1^ and has, to date, caused, approximately 412 million confirmed cases and about 5.8 million deaths^2^. Summarized under the name ‘COVID-19’ it elicits numerous symptoms like dry cough, fever, headaches, pneumonia, diarrhea, anosmia and others^3^ that are especially dangerous for the elderly and individuals with comorbidities^4^. Patients experiencing these symptoms, but also asymptomatic individuals (incidence ranging from 17.9 to 39.8%^5,6^), whose timeframe of viral shedding can even exceed the symptomatic carrier’s^7^, substantially contribute to transmission^4,8–10^ via respiratory droplets, fomites, direct or indirect contact^11^. Identification of these individuals via RT-qPCR, however, is difficult since the method is not necessarily suited for high-throughput analysis and difficult to implement in resource-limited environments^12^. Other drawbacks are comparability between labs^13,14^, an overestimation of infectious virions^15^, and a rather lengthy sample processing time of approximately 4-6 hours.

Hence, in recent years we have witnessed the emergence of a multitude of novel methods for the detection of viral nucleic acids, many of them employing CRISPR-effectors in combination with an amplification strategy for the genetic targets^16–18^. CRISPR, which is short for clustered regularly interspaced short palindromic repeats, was discovered as a mechanism of adaptive immunity in bacteria against viruses^19^ and has since been used in various applications in biotechnology – most prominently gene editing^20^ – in association with its effector: CRISPR associated protein 9 (Cas9). However, cleavage capability of the dual-RNA-guided endonuclease is restricted to double stranded DNA and requires the presence of a protospacer adjacent motif (PAM), located next to the target sequence^21^, which renders the effector unsuitable for the detection of circulating RNAs.

In this study, we extend the CRISPR-Cas-powered assay, that we already established for a signature microRNA (miRNA)-cluster, dysregulated in pediatric medulloblastoma patients^22^ onto a novel multiplexed, microfluidic electrochemical biosensing device, for the simultaneous detection of two genetic sequences characteristic for SARS-CoV-2. Instead of using Cas12 (a class 2 effector, targeting single-stranded DNAs and RNAs and upon activation by target recognition collaterally cleaving double stranded DNA^23,24^) like Broughton^16^, Ding^25^, or Gong and colleagues^26^, in this study we employ the Leptotrichia buccalis (Lbu)Cas13a effector and thereby bypass the need for a protospacer adjacent motif (PAM) in the target for recognition by the effector. Like for Cas12, target-specificity of LbuCas13a depends on guidance by a CRISPR (cr)-RNA^27^, which can be modified to target any RNA sequence of interest and is therefore perfectly suited to keep pace with spontaneous mutations in the viral genome^28^. Using LbuCas13a also circumvents reverse transcription, needed for both DETECTR and SHERLOCK^29,30^, of the viral genome. Employing the oligonucleotide sequences selected by the Charité, Institute of Virology (Berlin, Germany) we focus on one characteristic sequence within the envelope (E) and another within the RNA-dependent RNA Polymerase (RdRP) gene^31^, with the primer-probe set for the former being validated by Vogels and colleagues^32^.

Impairment of the immune system due to infection with SARS-CoV-2 will, in some cases lead to bacterial co- or superinfection, which is often treated with empiric antibiotic therapy, as a preventive strategy^33–35^. The success of the antibiotherapy, however, strongly depends on keeping the blood antibiotic concentrations within the “personalized” therapeutic range to minimize the risk of antibiotic resistance. Therefore, it is necessary to understand how the patient’s body metabolizes the drug with respect to the individual pharmacokinetic (PK) response^36^, i.e., to monitor antibiotic level fluctuations in the serum of a patient^37^. Here we introduce BiosensorX, a novel, multiplexed single-channel biosensor, onto which we immobilize the CRISPR-powered assay as well as a protein-based antibiotic detection assay **(Fig. 1)**. We thereby demonstrate the versatility of the biosensor’s design and provide a multitude of options and combinations of diagnostics, with respect to biomolecules of interest. We were able to conduct screening, confirmatory and discriminatory tests for SARS-CoV-2, as well as a control assay, simultaneously for two clinical specimens. Finally, we tested the credit-card format NFC potentiostat and microperistaltic pump as a compact measurement setup, further improving the point-of-care applicability of our system.

**Fig. 1.**
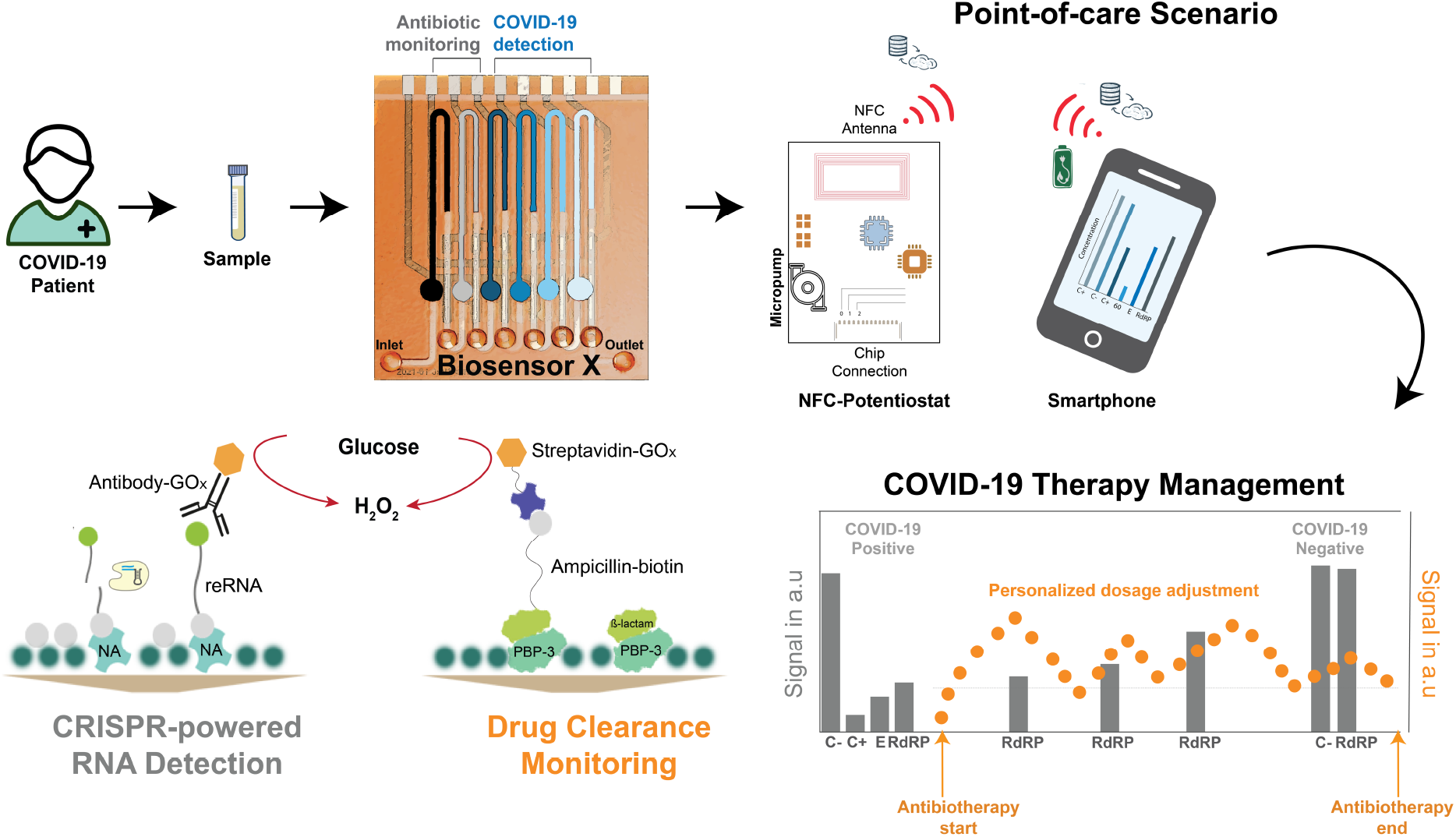
The proposed multiplexed microfluidic biosensor (BiosensorX) within the envisioned POC scenario, monitoring the treatment of bacterial co- or superinfection in COVID-19 patients. BiosensorX is capable of harboring both, the CRISPR-powered assays for the detection of multiple COVID-19-specific RNA sequences and a protein-based assay for ß-lactam monitoring. On-site patient monitoring could, hence, be performed by measuring antibiotic concentrations for personalized drug adjustment and screening for the abundance of the viral RdRP gene, to determine the end of isolation.

## Results

### Optimization of the CRISPR-powered RNA detection assay

For the simultaneous detection of multiple RNA sequences, we modified our existing bioassay^22^ to achieve lower detection limits as well as faster turnaround times: i) streptavidin-preincubation was substituted with neutravidin for its superiority in biotin-binding affinity **(SI Fig. 1a)**, ii) bovine serum albumin, previously used to prevent unspecific binding of biomolecules to neutravidin was substituted with casein: the smaller molecule is able to achieve satisfactory blocking in 30 minutes instead of 1 hour and reduces steric hindrance within the microfluidic channel, facilitating access for the reRNA’s biotin label to the binding sites of neutravidin **(SI Fig. 1b)**. Interestingly, when evaluated for neutravidin-biotin binding specificity the model assay failed to meet our requirements: in the absence of reRNA we were still able to observe a substantial, unspecifically generated amperometric signal **(SI Fig. 2a)**. iii) This background signal, however, could be quenched entirely by blocking the neutravidin binding sites, that were still open after application of the reRNA, by an additional application of biotin **(SI Fig. 2b)**. To test the improved assay’s performance and validate the crRNA and synthetic target oligonucleotides for the SARS-CoV-2 RdRP gene, we successfully conducted a proof-of-principle calibration (target concentrations of 1 nM, 100, 10 and 1 pM), using the standard incubation settings of 3 hours at 37 °C **(SI Fig. 3)**.

Further optimization of the assay, predominantly with respect to its limit of detection (LOD), but also concerning the variability of the individual measurements, was achieved by combining and switching combinations of two different Cas13a effectors (Leptotrichia wadeii (LwCas13a) and Leptotrichia buccalis (LbuCas13a)) with either a 14 nt-long reRNA (random sequence) or a reRNA consisting only of 20 uracil repeats (reRNA_20U)^38^ **(Fig. 2b-d)**. We observed the bioassay’s strongest performance when combining LbuCas13a with reRNA_20U. This was apparent in the sharp drop in the amperometric signal measured for 1 pM of target RNA (from undetectable levels to 104.19 µA cm^-2^; 30.4% of the blank signal). Consequently, all following experiments were performed using LbuCas13a as effector and reRNA_20U as the reporter of choice.

**Fig. 2.**
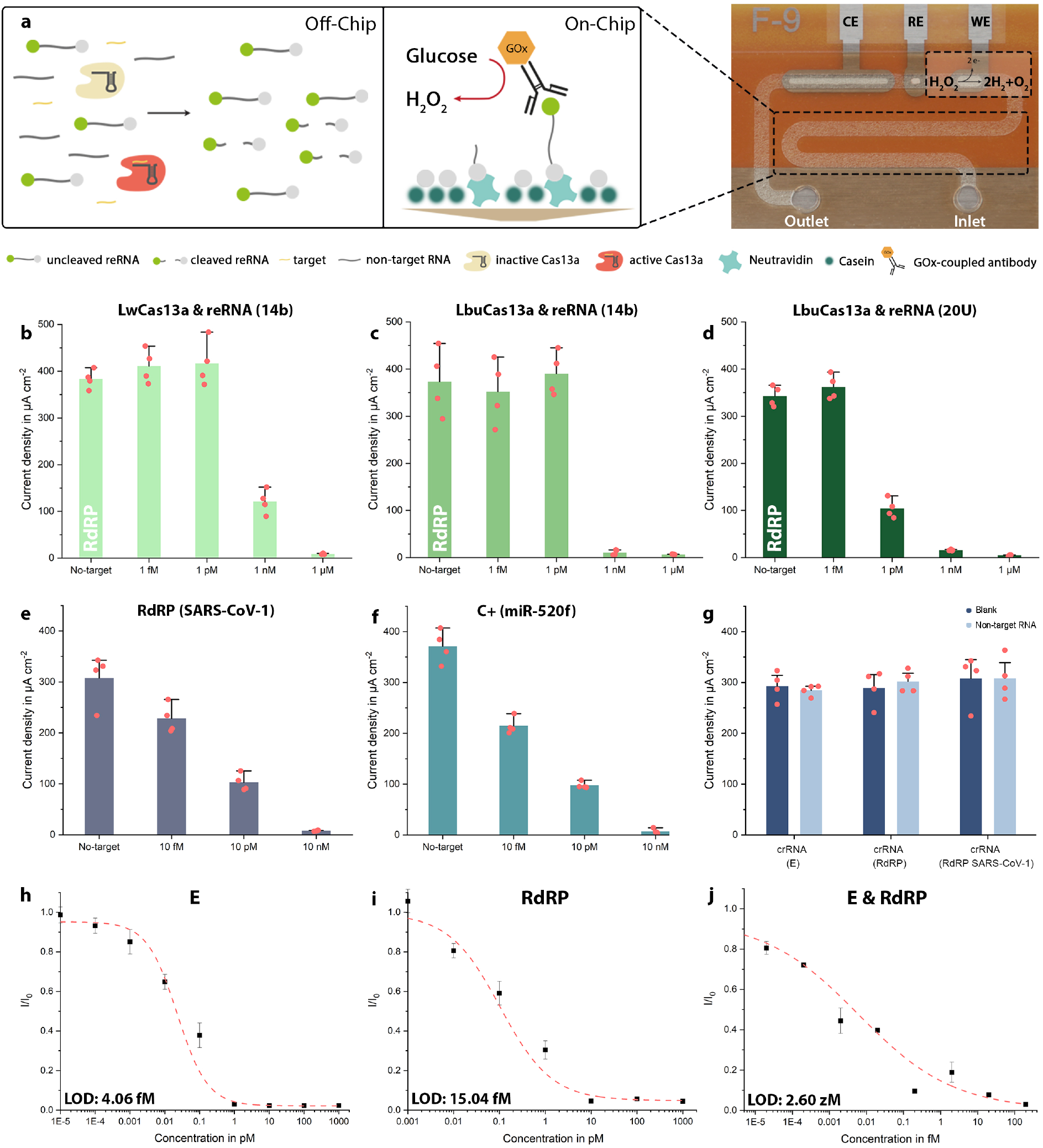
Optimization and calibration of target-amplification-free CRISPR-powered COVID-19 diagnostics. **a)** Schematic of the CRISPR-Cas13a-powered assay. LbuCas13a (beige: inactive; red: active) and crRNA form a complex within the sample solution, which also contains the target RNA (yellow) and reRNAs (labelled with a biotin (grey) and a 6-FAM molecule (green) on the 3’ and 5’-end, respectively). Upon activation by target recognition, the effector collaterally trans-cleaves the reRNA. The sample solution is then applied to the microchannel, which has been pre-functionalized with neutravidin (mint) and subsequently blocked with casein (dark green) to prevent unspecific binding of biomolecules to the channel’s surface. Subsequently, residual open binding sites are blocked by application of additional biotin. After immobilization of the assay, the GOx (orange)-conjugated anti-6-FAM antibody is introduced to enzymatically convert the glucose substrate to hydrogen peroxide. The redox species is then flushed over the electrochemical cell (WE, RE, CE) where it oxidizes at the WE. **b – d)** Show the comparison of the LwCas13a and LbuCas13a effectors in combination with either reRNA_14b or reRNA_20U. The strongest difference in signal amplitude could be achieved for 1 pM of target when using a combination of LbuCas13a with reRNA_20U, which was then used for all further experiments. **e)** Proof-of-principle measurements for SARS-CoV-1 RdRP (the negative control C-). **f)** Proof-of-principle measurements for miR-520f (C+). **g)** Cross-reactivity test, confirming the specificity of the different crRNAs to only their designated targets, respectively when tested with 100 pM of non-target RNA (crRNA (E): RdRP + C-; crRNA (RdRP): E + C-; crRNA (C-): RdRP + E). **h-j)** Sensor calibration for the synthetic SARS-CoV-2 E and RdRP gene, as well as a combination of the two, (crRNA: CRISPR-RNA, reRNA: reporter RNA, 6-FAM: 6-Carboxyfluorescein, GOx: glucose-oxidase, WE: working electrode, RE: reference electrode, CE: counter electrode). N = 4, error bars represent **+**SD.

### Positive and negative controls

Since RNAs are susceptible to degradation by omnipresent RNases, detection systems targeting these oligonucleotides are prone to produce false negative results. In sensitive settings, like nursing homes, hospitals or schools this can lead to disastrous results, with regard to the spread of viral material. It is therefore of utmost importance to ensure that each test is working correctly and to rule out contamination with RNases. We, therefore, implemented a positive as well as a negative control, which simultaneously serves as a discriminatory assay to rule out an infection with SARS-CoV-1^31^. The latter constitutes the measurement of a patient sample using a crRNA designed to target the RdRP gene, characteristic to SARS-CoV-1. If this measurement results in a signal of the maximum amplitude, we conclude that (i) the subject is not infected with SARS-CoV-1 and (ii) that none of the reagents are RNase contaminated. The positive control speaks to the general performance of the bioassay: in this case it employs a crRNA designed to target synthetic microRNA-520f (miR-520f, associated with proliferation of lung and gastric cancer cells^39,40^), which is added at a concentration high enough to completely eliminate the amperometric signal. These oligonucleotides could, however, be replaced by any other experimentally validated crRNA-target-pair, with a known amperometric readout that can be correlated to the utilized concentrations. To validate our negative and positive control oligonucleotide pairs we conducted measurements for 10 nM, 10 pM, 10 fM as well as a blank measurement (no-target) of the SARS-CoV-1 RdRP gene **(Fig. 2e)** and miR-520f **(Fig. 2f)**.

### Validation of crRNA specificity

To rule out the risk of false positive results produced by cross reactivity between crRNAs and non-target RNA, we measured the blank signals for each crRNA and compared them to the signal measured, when each of the three crRNAs (E, RdRP and SARS-CoV-1 RdRP, which, for ease of legibility, will further be referred to as C-) was incubated with both of the target RNAs (total concentration 100 pM, which would correspond to unrealistically high (∼500 million copies/µl) viral loads) for the other two crRNAs (crRNA(E): RdRP & C-; crRNA(RdRP): E & C-; crRNA(C-): RdRP & E). The non-target RNAs were not able to activate the effector and therefore no decrease in signal amplitude could be observed **(Fig. 2g)**. From this we conclude the target specificity of our crRNAs and thereby of the bioassay.

### Calibration of the sensor

In order to determine the statistical LOD of the biosensor, we performed measurements for different concentrations of the synthetic viral E and RdRP genes. Isothermal incubation (37 °C) of the sample solution for 5 minutes already provided results that were clearly distinguishable from the blank signal with the sensor reaching its LOD at 4.06 fM for the E and at 15.04 fM for the RdRP gene **(Fig. 2h-i)**. These target concentrations roughly correspond to 2,000 copies/µl (E) and 7,500 copies/µl (RdRP), and thereby supersede the LODs necessary to detect viral RNA in nasopharyngeal swabs (10^6^-10^9^ RNAs /swab)^41^ without target amplification. Calibrations measured on actual patient samples will, however, always fall short of the ones performed with synthetic RNA^42^. One solution proposed by Fozouni and colleagues, is a combination of crRNAs: Increasing the number of activated Cas13-effectors and thereby enhancing sensitivity while also providing a safeguard against mutational escape of the virus from detection^43^. We could verify this by combining crRNA(E) with crRNA(RdRP) to record an additional calibration curve, using the synthetic sequences as targets. The analytical LOD of this approach reached 2.6 zM **(Fig. 2j)**. Another way of circumventing the diminished LOD, obtained when measuring actual samples instead of synthetic targets, is prolonging the reRNA-cleavage time. After even a slight increase from 5 to 10 minutes, we were able to observe a decreased signal amplitude from 66.05% to 42.36% of the blank signal for a target concentration of 100 fM (50,000 copies/µl). Accordingly, after an incubation period of 2 hours, the amperometric signal had decreased to 3.08 % of the blank signal **(SI Fig. 4)**.

### Implementation of the single-channel multiplexed BiosensorX

In an attempt to provide a single platform that can perform both measurement of antibiotic concentration and detection of viral RNA in clinical samples, we combined two different assays on a 6-fold BiosensorX **(Fig. 3a)**: (i) a protein-based assay for ß-lactam monitoring^37^ and (ii) a CRISPR/Cas13a powered assay for RNA sequences characteristic to an infection with COVID-19 **(Fig. 3b)**. To mimic the drug clearance behavior, the first two immobilization areas of BiosensorX were used to perform antibiotic measurements for simulated dosages of piperacillin/tazobactam along with a negative control (blank signal: no antibiotic present in the sample). The three measurement time points represented the start (t = 0, no antibiotic treatment yet), the initial phase (t = 5 min, high levels of antibiotic) and the clearance (4 hours after the initial dose) of the drug within treatment **(Fig. 3c-f)**.

**Fig. 3.**
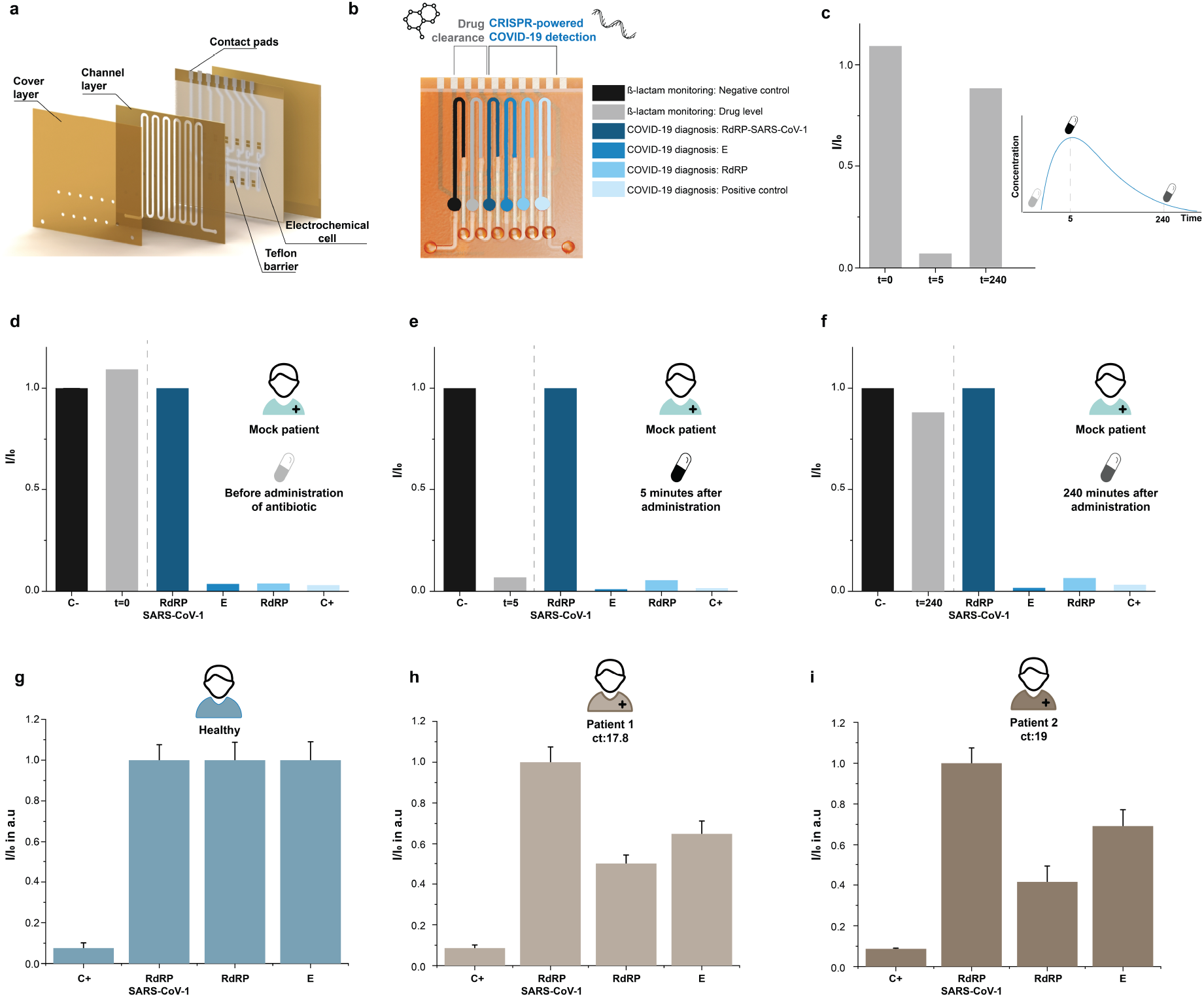
Multianalyte detection capability of BiosensorX. **a)** 3D rendering of the stacked BiosensorX. **b-f) Personalized antibiotic dosing in a mock COVID-19 patient. b)** Methodology of simultaneous measurement of antibiotic concentration and detection of viral RNA in clinical samples. **c)** Overview of the drug clearance behavior in a mock patient infected with SARS-CoV-2 **d)** before, **e)** 5 minutes, **f)** 240 minutes after “antibiotic administration” (n = 1). **g-i) Clinical samples from a healthy subject and two COVID-19 patients analyzed with BiosensorX**. Current densities for **g)** a healthy subject, **h)** a patient with Ct 17.8 and **i)** a patient with Ct 19. Signals obtained from the samples of diseased patients were normalized to their respective target-equivalents obtained from the healthy subject and are therefore presented in arbitrary units. N = 2, Error bars represent +SD.

When the drug concentration was measured 5 minutes after administration, we could observe a sudden drop in the measured signal, indicating a sharp increase in the ß-lactam level. At t = 240 min, the drug concentration is assumed to decrease towards the control level, which we were also successfully able to measure on the multiplexed sensor. Measurement of the viral load (indicated by the respective abundance of E and RdRP copies) is not expected to change within the observed timeframe and is therefore simulated to stay at the initially (t = 0) measured 1 pM (∼500,000 copies/µl) within this experiment. The negative control for the CRISPR-based assay in this case was performed with 5 µl of RNase free PBS (10 mM). The positive control successfully detected 1 nM of spiked-in miR-19b, a biomarker for pediatric medulloblastoma, validated in a previous study from our group^22^. These results indicate that the proposed multiplexed biosensor is capable of harboring multiple different bioassays from various biomolecule classes without cross-contamination.

### Measurement of clinical specimen

To test the applicability of our multiplexed platform for SARS-CoV-2 detection, we analyzed nasopharyngeal swab samples of two COVID-19 patients with high viral loads (Ct 19 and 17.8, respectively) as well as one specimen taken from a healthy subject. We employed the 4-fold BiosensorX to conduct the positive control (in this case miR-520f; 100 pM), and discriminatory (RdRP SARS-CoV-1) assay, as well as the screening (E) and confirmatory assay (RdRP SARS-CoV-2) simultaneously (**Fig. 3g-i**). Each sample was measured on two individual multiplexed biosensors and the signals obtained from the infected individuals for the E and RdRP genes were normalized to their counterparts measured for the healthy subject. We were able to obtain consistent values for C+ (15.82 ± 3.66 nA) as well as the discriminatory assay (210.15 ± 7.64 nA) across all subjects and observed a considerable drop in the signals measured for the RdRP gene in both patients (patient 1 (Ct 17.8): 41.52% and patient 2 (Ct 19): 50%). A clear, decrease in the E gene signal was also visible for both patients (patient 1 (Ct 17.8): 69.18% and patient 2 (Ct 19): 64.87%).

## DISCUSSION

Over two years of the still ongoing COVID-19 pandemic has lead researchers to the consensus that the frequency and sample-to-result time of tests should be prioritized over an LOD that can compete with that of RT-qPCR (LOD: 10-100 copies/µl, full range of viral titers between Ct 17-38 and turnaround times of 24-48 hours)^15,32,44,45^. Peak viremia has been reported at the time of symptom onset with infectiousness declining until day eight to ten after which the probability of transmission is almost none. In some cases, positive PCR-results can be obtained until as much as 50 days after onset, at which point the patient is very unlikely to still be infectious^41,46,47^. Generally, an individual is considered infectious if both the antigen as well as the PCR tests show positive results, whereas viral loads below 10^3^ copies/µl are associated with a low risk of infectiousness^41,48^.

While RT-qPCR is still considered the gold standard for the detection of nucleic acids, quantitatively comparing different Ct levels across laboratories is difficult (some studies describe Ct values <30 as ‘high’^48^, for others Ct values >24 are already classified as ‘low’^49^ with again other studies reporting plaque forming units only for Ct 13-17 progressively decreasing until Ct 33^50^). Also unbalanced amplification of short subgenomic sequences in poor clinical samples can lead to an overestimation of the viral load^51^ and varying waiting times (average in Germany fluctuating between 8-48 hours^52^, while the US reported three to ten days^53^) can undermine the effectiveness of the test^54^. Hence, there has been a plethora of methods developed with the intention to optimize testing strategies for the broad population and substitute RT-qPCR with its expensive equipment and requirements for specifically trained operators.

The most common form of COVID-19 diagnostic to date is the rapid antigen test on a lateral flow device (LFD). Due to their easy handling, short sample-to-result time and straightforward readout, test kits employing LFDs have been established for at-home testing, as well as for state-certified tests (Germany), with sensitivities between 70-80% (>90% sensitivity for patients with Ct<25) and specificities ≥99.7%^55,56^. Results are usually provided within 30 minutes, but also require high viral loads to turn out positive^57^, which begs the question: Are people tested negative with an LFD today going to be infectious tomorrow? I.e., have they been exposed to the virus but have not yet amplified the virus particles to supersede the LOD of the test?

An approach to circumvent this issue is the utilization of CRISPR/Cas systems, which, even without amplification strategies are able to reach LODs below 50 fM (∼30,000 copies/µl)^58,59^ and can be paired with various signal readout options (fluorescence, colorimetric lateral flow, etc.). Examples for the CRISPR-effectors employed in COVID-19 tests so far are Cas3, Cas12a/b, Cas13a and the effector used in this study, Cas13a. The latter can achieve remarkably low LODs due to its capability of trans-cleaving approximately 10^4^ (re)RNA molecules in its vicinity upon activation by recognition of the target sequence^60^. Nevertheless, most methods developed for the detection of SARS-CoV-2 genes employ either reverse transcription loop-mediated isothermal amplification (RT-LAMP; LOD: 100 copies/µl, 90 min sample-to-result time^61^) or reverse transcription recombinase polymerase amplification (RT-RPA; LOD: 2.5-100 copies/µl, sample-to-result-time: 20 min - 1 hour^42,62,63^) as a target-amplification strategy or even both (LOD: 82 copies/µl after ca. 50 min^64^). Another way of lowering the LOD, proposed by Fozouni and colleagues, is the combination of multiple target-complementary sequences into a single crRNA (∼200 copies/µl in under 30 min)^65^.

In this study, we combine a multiplexed CRISPR-based detection system with an electrochemical readout to target unamplified oligonucleotide sequences characteristic for SARS-CoV-2: one within the envelope (E) and another from the RNA-dependent RNA Polymerase (RdRP) gene^31^, with the primer-probe set for the former being validated by Vogels and colleagues^32^. The former non-specifically indicates infection with a member of the Betacoronavirus genus^66^, the latter is specific to SARS-CoV-2^31^. Like Corman and colleagues we also implement a discrimination assay to rule-out infection with the 2003 SARS-CoV, using a characteristic target within the SARS-CoV-1 RdRP gene. While this method requires slightly more expertise than the optical readout of a conventional LFD, it only produces a fraction of the waste. In addition, the substrate, glucose, is stable over long periods of time, inexpensive and non-toxic. Within a sample-to-result time of about 30 min we were able to obtain an LOD of 2,000 copies/µl for the E gene, 7,520 copies/µl for RdRP and <1 copy/µl for a combination of the targets. However, since all calibrations were measured using synthetic target-oligonucleotides, the LODs cannot be translated directly to clinical specimen, which are likely to be much higher, due to a multitude of different RNAs, other than only the reporter. An indication therefore are the proof-of-principle measurements that we conducted on the clinical samples of two COVID-19 patients with high viral loads. We could, however, measure distinctly reduced amperometric signals for the RdRP gene in these samples, whereas interpretation of the results for the E gene was difficult. Because of the discrepancy in their respective LODs (E: 4.06 fM; RdRP: 15.04 fM) and also because E (as well as Orf7 and the nucleocapsid gene) is highly abundant in subgenomic RNA, which often leads to a 100-1,000-fold overestimation of a patient’s viral load^51^ the E gene signals were assumed to fall below the ones measured for RdRP, which they did not. One possible explanation for these results is a mutation within the E gene sequence of the viral genome that occurred at some point since the establishment of the Corman protocol in 2020, leading to partial failure of recognition by the crRNA, that was designed according to a protocol from 2020^31^.

Despite the relatively short turnaround time, adequate LOD and reasonable pricing (3 € for the entire COVID-19 diagnostics on a 4-fold biosensor; see **Supplementary Table S3**) of our proposed biosensor, there is still potential for improvements in the meaningfulness of its results and point-of-care applicability. First, a combination of viral RNA detection with serological assays could increase its informative value by correlation of the viral load to the type of antibody that is expressed at the time of sample acquisition^67,68^, as proposed by Puig and colleagues^69^. Another possibility is a combination with the detection of virus as well as host miRNAs, expressed during an infection with COVID-19, since these can influence disease progression and lead to vastly different outcomes in patients^70^. An expansion of RNA-targets to either these miRNAs or new variants of SARS-CoV-2^71^ could facilitate the development of specified treatment plans. Studies have also shown that the Cas13a-crRNA complex is stable^58^ and can be lyophilized and rehydrated for use at the POC, which would improve its applicability in remote locations to which continuously refrigerated transport is difficult. In addition, our experimental results could also benefit from optimization of the upstream sample processing. While we avoid the risk of skewing our analysis by unbalanced amplification of RNAs, sample preparation is still a factor that can have major impacts on the experimental outcome. Choice of the lysis buffers, for example, can yield vastly variable results, depending on its components^42^ and reported success of RNA-extraction versus no-RNA-extraction results are contradictory. Untreated nasopharyngeal samples are likely to contain RNases^61^ that would need to be inactivated prior to lysis and while Myhrvold and colleagues developed a nuclease inactivation and viral lysis method (HUDSON) to omit an extra RNA extraction step^72^ and Ladha and co-workers condensed their isolation protocol down to a 5-min procedure^73^ other studies claim the sample processing step to be unnecessary^12,74,75^. Furthermore, as there are still 5 steps that require user interventions, our protocol is prone to RNase contaminations and errors due to misuse. Reducing these steps to ideally only three hands-on steps, (addition of the sample to the mastermix, application of the mastermix to the microfluidic device and readout) requires further studies. These are, however, warranted, since, due to constantly evolving new variants, but also waning of the neutralizing antibody response^76,77^ the need for rapid and frequent testing for SARS-CoV-2 will likely remain.

The platform introduced in this study presents a direct approach to quantifying the viral load of a clinical sample in approximately 30 min with the potential for combination with a variety of assays from other biomolecule classes. In this case, we chose protein-based monitoring of antibiotic levels, since bacterial co- or superinfections often coincide with viral primary infections and are preventatively treated with generic antibiotics. The obtained amperometric signal directly correlates to viral copy numbers and may be used for quantification of viral loads across laboratories, without the need for extensive system calibrations. Since we use a syringe pump, an in-house designed chip-holder with attached printed circuit board and a potentiostat, our method is suited for testing facilities, e.g., in retirement homes, schools, or prisons where it could be used for rapid and frequent screening for infectious individuals. Additionally, we were able to miniaturize the on-site measurement setup by using a microperistaltic pump and a credit card-sized NFC potentiostat (**Fig. S13**) to improve the cost effectiveness and ease of setup. In conclusion, frequency of screening is understandably favored in the trade-off with accuracy, it is advisable to work with all three: high frequency, low LOD and short sample-to-result / waiting times, as well as to educate about the risks and benefits of vaccinations to limit misinformation and to contain the spread of SARS-CoV-2.

## Supporting information

Statement of ethic commission

## Data Availability

All data produced in the present study are available upon reasonable request to the authors.

## Acknowledgements

The authors would like to thank the Deutsche Forschungsgemeinschaft (DFG, German Research Foundation) and Bundesministerium für Bildung und Forschung (BMBF, Federal Ministry of Education and Research) for partially funding this work under grant numbers 404478562, 421356369, 390939984 (EXC 2189 – CIBSS), and 13GW0493 (MERGE). The authors also thank Nadine Urban, Dr. Julia Baaske and Dr. Barbara Enderle for providing organizational assistance and technical advice, respectively. We also would like to acknowledge Jobst Technologies GmbH (an iST AG company – Freiburg, Germany) and Silicon Craft Technology PLC (Bangkok, Thailand) for providing us with a microperistaltic pump (MP.CPP1.180.ZM) and an NFC potentiostat (SIC4341, potentiometric sensor interface with NFC type 2), respectively.

## Methods

### Electrochemical Microfluidic Biosensor

The dry-film-photoresist-based biosensing devices used in this study (2×0.8 cm^2^) are manufactured according to our previous work^78^ and house two individual biosensors. In brief, each sensor features a microfluidic channel, which is subdivided into two functional units: the immobilization area (length: 25 mm, surface area: 10.34 cm^2^), onto which all components of the bioassay are immobilized and the electrochemical cell, containing a working electrode (platinum, Pt), a reference electrode (silver/silver chloride, Ag/AgCl) and a counter electrode (Pt). The two units are separated by a hydrophobic stopping barrier, facilitating the containment of fluids (0.58 µl), which are applied to the channel by capillary forces (**Fig. 2a**). Similar to the single-analyte version, the multiplexed electrochemical biosensor consists of either four (0.61 µl) or six (0.67 µl) functionalized zones (length: 19 mm, surface area: 12.27 cm^2^ for the four-channel and length: 19 mm, surface area: 12.93 cm^2^ for the six-channel version), each complete with their own inlets, outlets and electrochemical cells. With this design, it is possible to functionalize each immobilization area separately which enables a combination of different assays on the same chip. While the electrochemical cells of single and 4-analyte designs have their own counter, reference and working electrode; in the 6-analyte design, all electrochemical cells have one common reference and counter electrode, and an individual working electrode.

### Production and Purification of Lbu-C2c2 (LbuCas13a)

Lbu-C2c2 bacterial expression vector (Addgene #83482) is transformed into *E. coli* BL21 (DE3) pLysS (Invitrogen). Bacteria are grown in LB medium (Carl Roth, Germany) containing 100 µg mL^−1^ ampicillin (Roth) and 36 µg ml^−1^ chloramphenicol (Carl Roth, Germany) at 37 °C, 150 rpm. Once OD_600_ = 0.6 is reached, protein production is induced with 1 mM isopropyl ß-D-1-thiogalactopyranoside (IPTG) (Carl Roth, Germany) for 16 h at 16 °C. Cells are harvested by centrifugation at 6,000 × g for 10 min and the bacterial pellets are resuspended in Ni-lysis buffer (50 mM NaH_2_PO_4_, 300 mM NaCl, 10 mM imidazole, pH 8.0) and lysed using a French press (APV 2000, APV Manufacturing, USA) at a maximum of 1,000 bar for three rounds. Cellular debris is removed by centrifugation at 30,000 × g for 30 min at 4 °C and the supernatant is purified by gravity flow Ni^2+^-NTA chromatography using superflow agarose (Qiagen GmbH, Germany) following the manufacturer’s instructions and the eluate is supplemented with 1 mM tris(2-carboxyethyl)phosphine (TCEP) (Carl Roth, Germany) and 15% (v/v) glycerol and concentrated using a Spin-X UF 6 (100k MWCO, Corning, USA) to a final concentration of 0.45 mg ml^−1^ of pure protein. The purified protein is analyzed by SDS-PAGE (12% (w/v) gels) and Coomassie brilliant blue staining. The protein concentration is determined by use of the Bradford method (Bio-Rad laboratories, USA) with bovine serum albumin (Carl Roth, Germany) as standard and the concentration of the pure protein is determined by analyzing the SDS gels with ImageJ. Aliquots are stored at –80 °C and freshly thawed for each experiment.

### CRISPR-Cas13a-powered Assay

All tests are performed on pre-incubated microfluidic biosensor chips: the immobilization areas are first incubated with neutravidin (0.8 mg ml^−1^ in PBS (10 mM; pH 7.4); 31000; ThermoFisher Scientific, USA) for one hour, followed by a 30-minute incubation with casein (85R-108; Fitzgerald, USA), both at 25 °C. These steps provide high affinity binding sites for the biotin label on the reporter RNA (reRNA_20U) and block unspecific binding of biomolecules to the surface of the microfluidic channel. The reaction mix, comprising LbuCas13a, a target-complementary CRISPR RNA (crRNA, Biomers GmbH, Germany), the reRNA labelled with biotin and 6-FAM on the 3’ and 5’-end, respectively (Reporter_RNA_20U_2’OMe-A, Biomers GmbH, Germany) and a murine RNase inhibitor (M0314L, New England BioLabs, Germany) (all in Tris-buffer (MgCl_2_ (6 mM), Tris (40 mM), NaCl (60 mM); pH 7.3)) is incubated for 10 minutes (37 °C). The targets, characteristic sequences from the SARS-CoV-2 E, RdRP as well as the SARS-CoV-1 RdRP gene (Biomers GmbH, Germany) are then added for a 5-min incubation at the same temperature (the synthetic target sequences, and their corresponding crRNAs were designed according to the diagnostic protocol of Charité Berlin^31^). Upon target recognition, the LbuCas13a-crRNA complex is activated and trans-cleaves all reRNAs in its surrounding. For information on oligonucleotides used in this study please refer to **Supporting Information**. The sample solution is then applied to the immobilization area for 1 minute (25 °C). In order to prepare the antibody-mediated detection, additional biotin molecules are required to saturate binding sites that are still open after application of the sample solution. As the glucose oxidase (GOx)-conjugated (see **Supporting Information**) monoclonal anti-6-FAM antibody (SAB4600050-125UL, Merck, Germany) will now only bind to the 6-FAM label on the 5’ end of uncleaved reRNA, the signal obtained in the amperometric readout is highly specific and inversely proportional to the target concentration within the sample solution.

### β-lactam Detection

Functionalization of the immobilization area is accomplished by incubation with penicillin binding protein 3 (PBP-3^37^; 250 µg ml^−1^ in PBS (10 mM; pH 7.4), expressed via in-house production) for 1 hour, followed by 30 minutes of blocking with biotin-free casein (85R-108, Fitzgerald, USA). The third step of the assay relies on the competitive binding of piperacillin-tazobactam and/or biotinylated ampicillin (0.2 µg ml^−1^ in PBS (10 mM; pH 7.4) obtained via in-house production) to PBP-3. At the last step, the channel surface is incubated with avidin-GOx (50 µg ml^−1^ (A4500-70.2, Biomol, Germany) in PBS (10 mM; pH 7.4) for 15 minutes.

Unbound biomolecules are flushed (PBS (10 mM) with 0.05% Tween, pH 7.4, P3563, Merck, Germany) after each incubation step for an accurate amperometric readout. Changing the incubation solutions as well as rinsing of the microchannel are performed by application of a vacuum to the outlet of the microchannel in order to avoid contamination of the electrochemical cell.

### Electrochemical Signal Readout

Once the immobilization of the assay has been completed the biosensor is placed into a custom-made chip-holder on a printed circuit board (Beta LAYOUT GmbH, Germany), connecting to a four-channel potentiostat (MultiEmStat3 for single and 4-analyte biosensor, EmStatMux8 for 6-analyte biosensor; PalmSens, The Netherlands). The inlet and outlet of the chip’s microchannel are then connected to a syringe pump (PHD2000; Harvard Apparatus, USA) via silicone tubes (DENE3100504; VWR, Darmstadt, Germany). The electrochemical readout of single- and 4-analyte biosensors is preceded by preconditioning of the working electrodes; the electrode’s potential is cycled (5 seconds at each 0.8 and –0.05 V vs. the on-chip reference electrode) 30 times, while PBS (10 mM, pH 7.4) is passed through the channel and over the electrochemical cell at a flow rate of 20 and 10 µl min^−1^, respectively. Preconditioning of the 6-analyte biosensors is performed with a 10-cycle cleaning (5 seconds at each 0.8 and –0.05 V vs. the on-chip reference electrode), while the flow rate of PBS (10 mM, pH 7.4) is 10 µl min^−1^. After this process, manufacturing residues are removed from the electrode surfaces. Amperometric signal detection is performed by applying a stop-flow-protocol to the flow of the substrate solution (glucose, 40 mM in PBS (10 mM; pH 7.4); Merck, Germany): filling the entire microchannel with the substrate (at a flow speed of 20 µl min^-1^ for single-analyte and 10 µl min^-1^ for multianalyte biosensor) is followed by a 2-minute delay, during which the immobilized GOx catalyzes the conversion of glucose into hydrogen peroxide. Upon restart of the flow the accumulated redox reporter is flushed over the electrochemical cell and oxidizes at the working electrode. During “flow” phase of multianalyte biosensor, these redox reporters are also passing through neighbouring electrochemical cells in addition to their own individual electrochemical cell, which creates the additional successive peaks. The digital readout of the amperometric signal^79^ as well as the pre-conditioning of the working electrodes are performed with the corresponding software (Multitrace 4.3 for single and 4-analyte biosensors, PSTrace for 6-analyte biosensor; PalmSens, The Netherlands).

### Clinical Samples and RNA-Extraction

Patient samples were provided by the Medical Center – University of Freiburg and lysed in the Alinity Lysis Solution (09N20-001, Abbott, USA). Subsequently, the RNA extraction was performed using RNeasy MinElute Cleanup Kit (Qiagen GmbH, Germany) according to manufacturer’s instructions in an RNAse-free environment. It needs to be noted, that due to very small volumes of isolated RNA the discriminatory assay for patient 2 was conducted with PBS instead of actual clinical sample.

### Miniaturization of the Measurement Setup for Point-of-care Testing

To perform the amperometric signal readout, the microfluidic biosensor needs to be electrically and fluidically integrated into the measurement setup via a custom-made chip holder^78^. The setup used in this study includes a syringe pump to realize a constant fluid flow through the biosensor, a potentiostat for signal readout and a laptop for the visualization of the measured signal. For an improved on-site patient monitoring or even self-testing, however, the equipment needs to be simplified and miniaturized. To respond to that need, we compared the fluidic control of our syringe pump (PHD ULTRA™ 4400, Harvard Apparatus, USA) to a microperistaltic pump (MP.CPP1.180.ZM, Jobst Technologies, Germany) to ensure the necessary constant flow rate during the stop-flow measurement. Subsequently, we evaluated the performance of the NFC-potentiostat by comparing the signal readout with that obtained using a bench-top potentiostat (**Figure S11 and S12**). Both measurements were successfully performed using a single-channel biosensor incubated with the model assay (100 µg ml^−1^ Avidin-GOx).

Replacing the bench-top potentiostat and syringe pump with an NFC-potentiostat and a microperistaltic pump, respectively, greatly contributes to the miniaturization of the measurement equipment as both powering of the potentiostat and the pump, as well as visualization of the measurement results could be achieved by a smartphone or other smart devices, instead of a laptop (**Figure S13**).

