## Supplementary Information for "Multiplexed biosensor for point-of-care COVID-19 monitoring: CRISPR-powered unamplified RNA diagnostics and protein-based therapeutic drug management"

**Model Assay**

**Table S1. Components of the model assay.** All biochemical compounds are diluted in 10 mM PBS (pH 7.4) and incubated at 25 °C.

| Compound | Concentration | Incubation Time |
| --- | --- | --- |
| Avidin/Streptavidin/Neutravidin | 800 µg ml^–1^ | 1 h |
| BSA/Casein | 10 mg ml^–1^/ undiluted | 1 h / 30min |
| reRNA | 250 mM | 5 min |
| 6-FAM antibody | 10 µg ml^–1^ | 5 min |

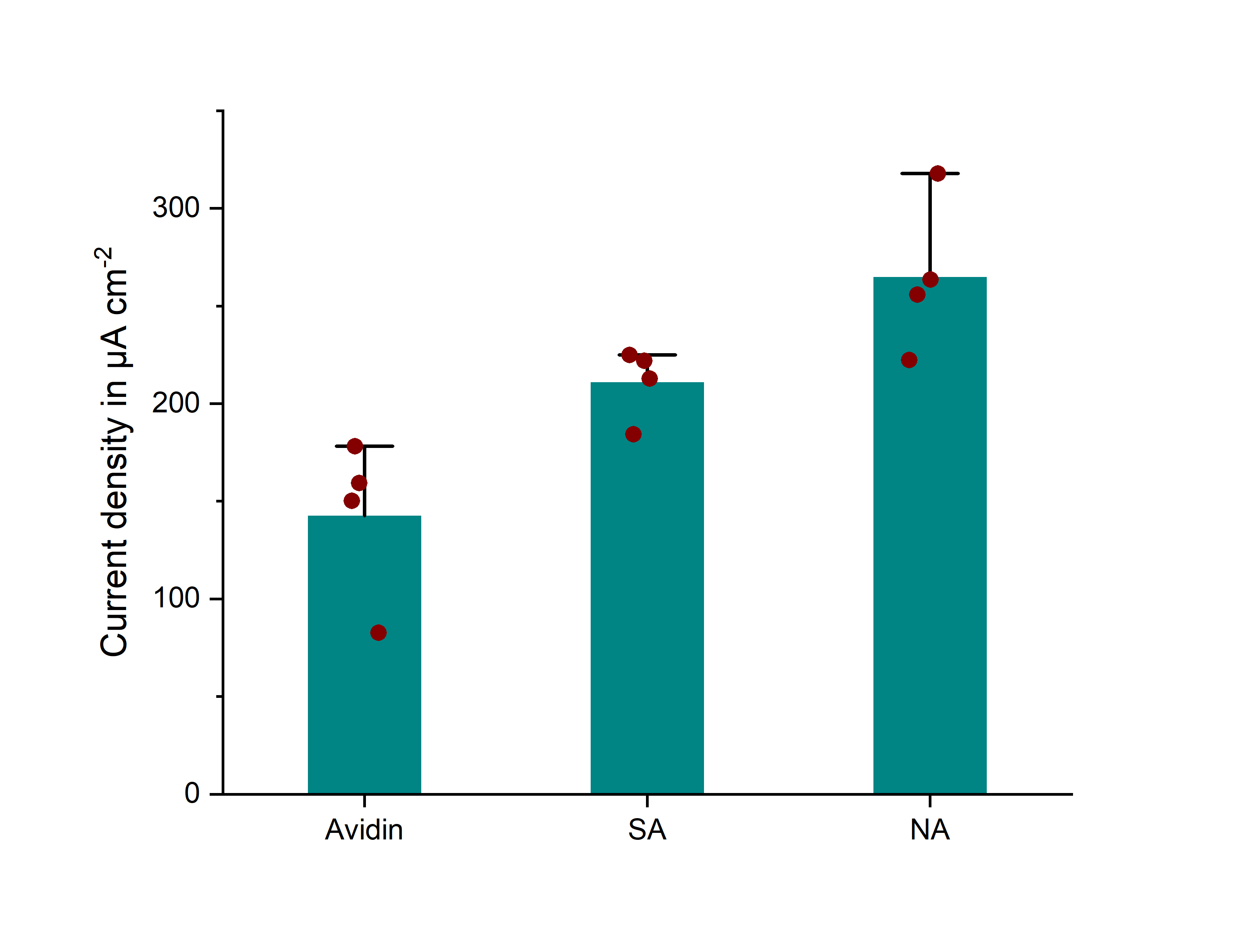
**Optimization of the CRISPR Assay**

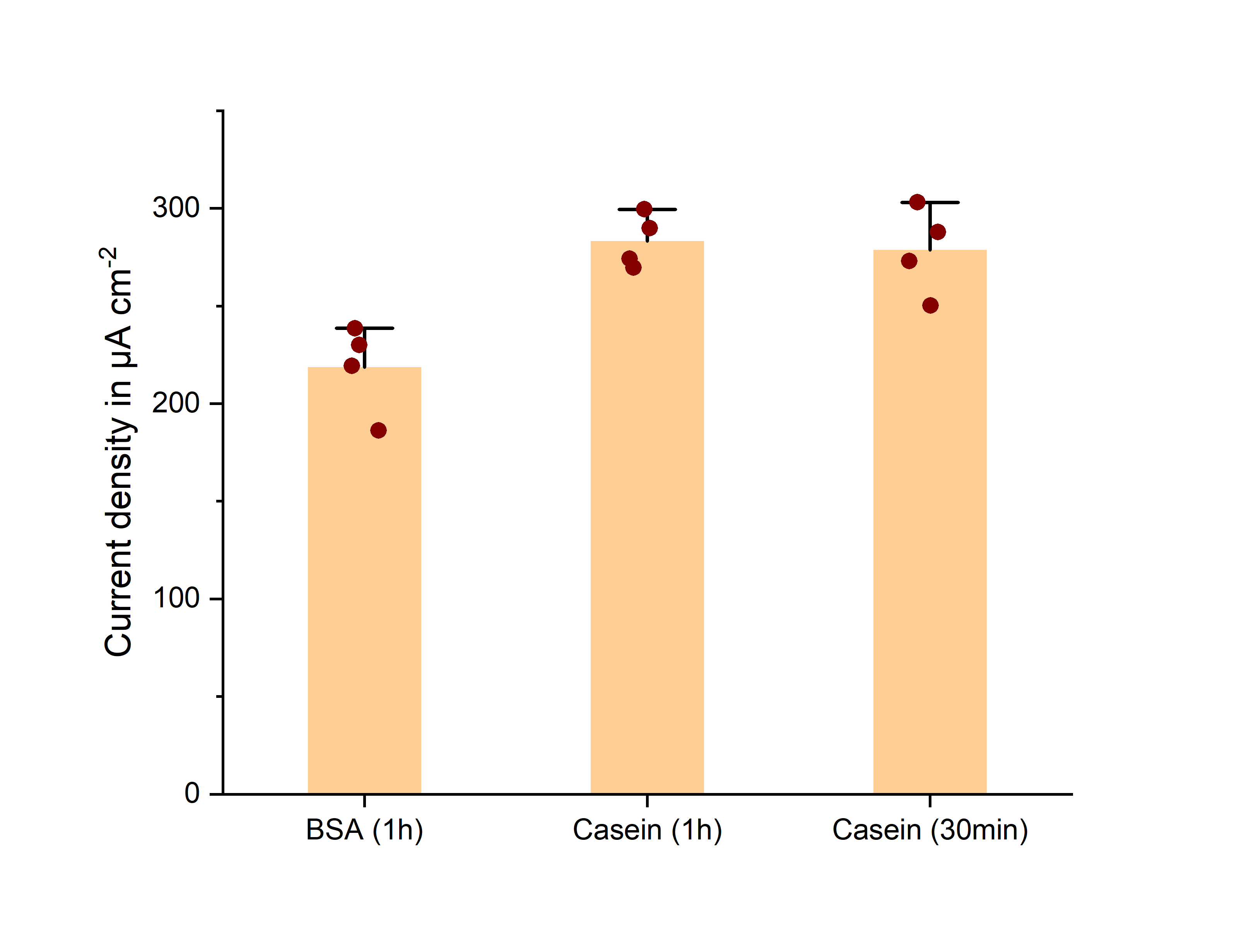

**a**

**b**

**Figure S1. a) Comparison of different avidin analogues for pre-functionalization of the microfluidic channel with the model assay**. All avidin analogues were applied at the same concentration (800 µg ml^–1^ in PBS 10 mM, pH 7.4), Neutravidin shows the strongest signal when tested using the model assay and is therefore used for all following experiments. **b)** **Comparison of different blocking strategies with the model assay**. All methods were tested at the same incubation temperature of 25 °C. Application of casein for 30 min shows an average signal height comparable to 1 hour of BSA incubation (as performed in previous studies). Error bars represent +SD of n = 4 (SA: Streptavidin, NA: Neutravidin BSA: Bovine Serum Albumin).

**Specificity Test**

**b**

**a**

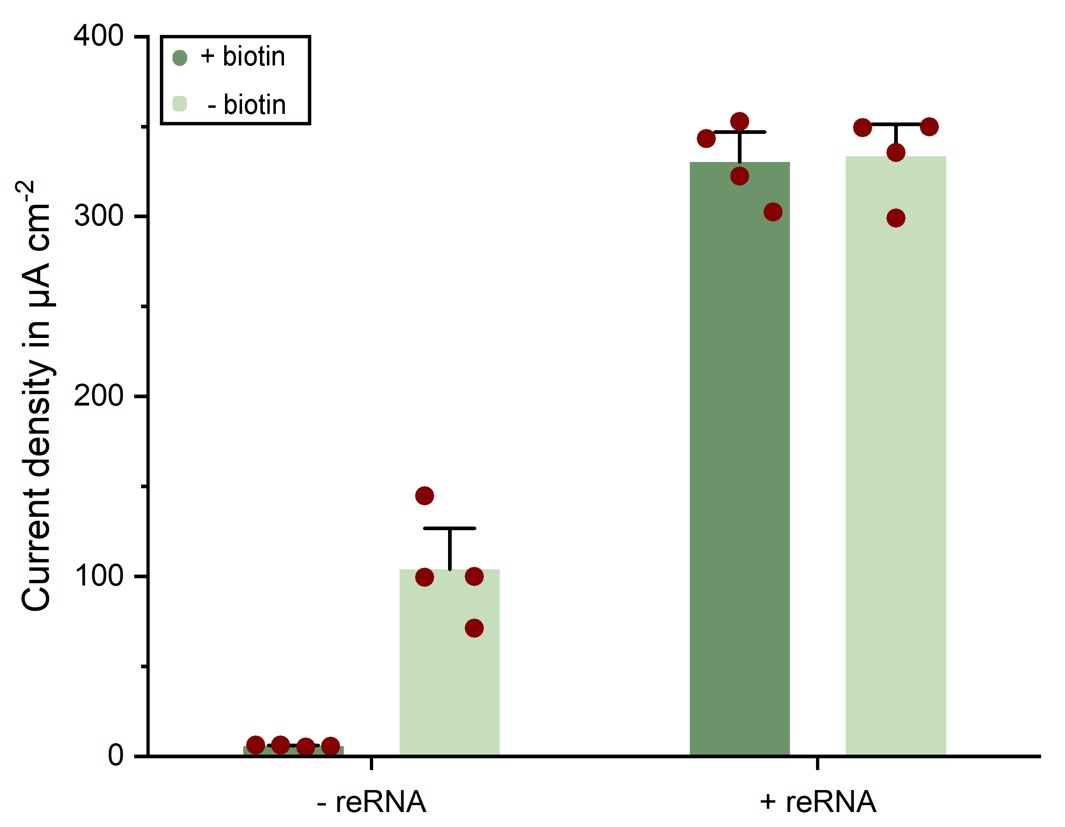

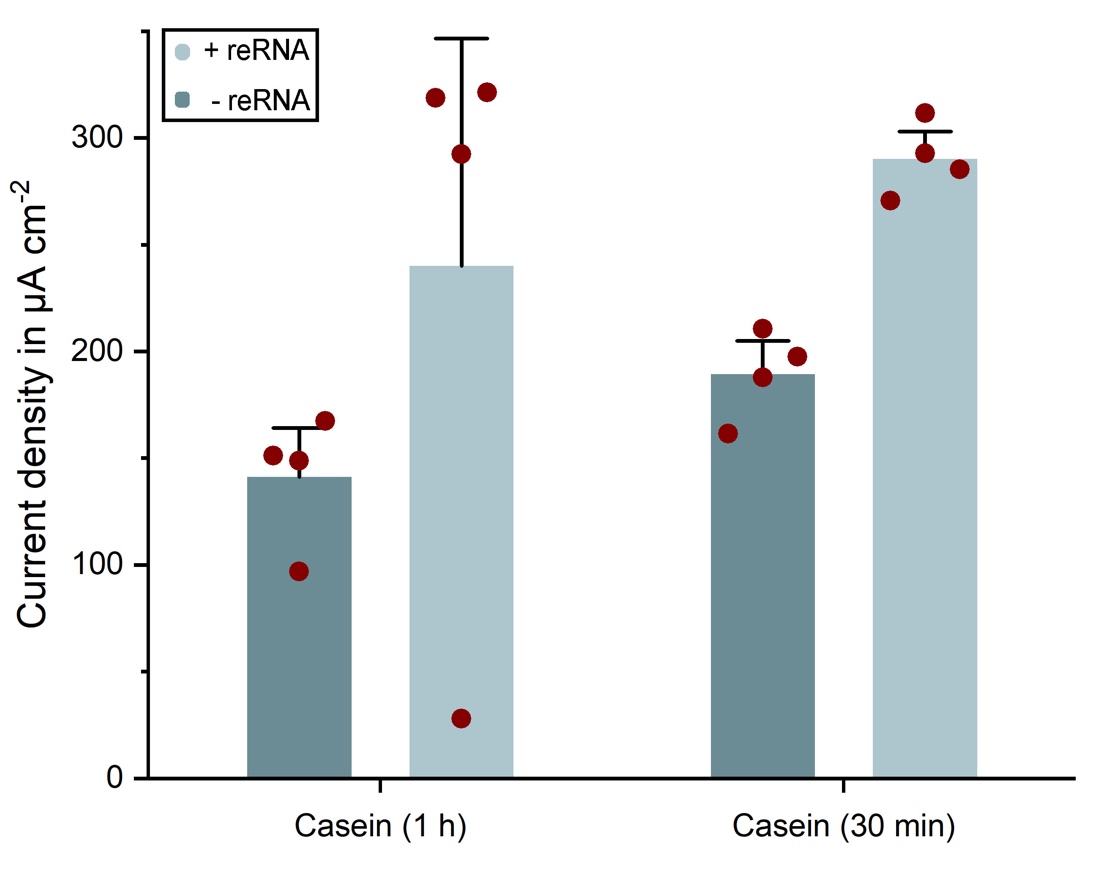

**Figure S2. a)** **Specificity test for neutravidin using the model assay**. Despite the casein blocking step neutravidin still exhibits strong unspecific binding activity towards the anti-6-FAM antibody, resulting in 141.12 µA cm^–2^ (1-hour casein incubation) or even 189.34 µA cm^–2^ (30-min casein incubation) in the absence of reRNA. **b)** **Application of biotin after reRNA incubation can quench unspecific signals in the model assay**. Signals were obtained after a 30-minute incubation with casein. When applied in the presence of reRNA the average signal is reduced to only the substrate that is generated by the glucose oxidase-conjugated antibodies that are in fact bound to the 6-FAM label on the reporter and not unspecifically to other biomolecules or the channel surface. This hypothesis is validated by the signals obtained from biotin post saturation in the absence of reRNA. Based on these results all following experiments were performed by blocking with casein for 30 min at 25 °C (Error bars represent +SD of n = 4 (reRNA: reporter RNA).

**Standard Off-Chip Cleavage Test**

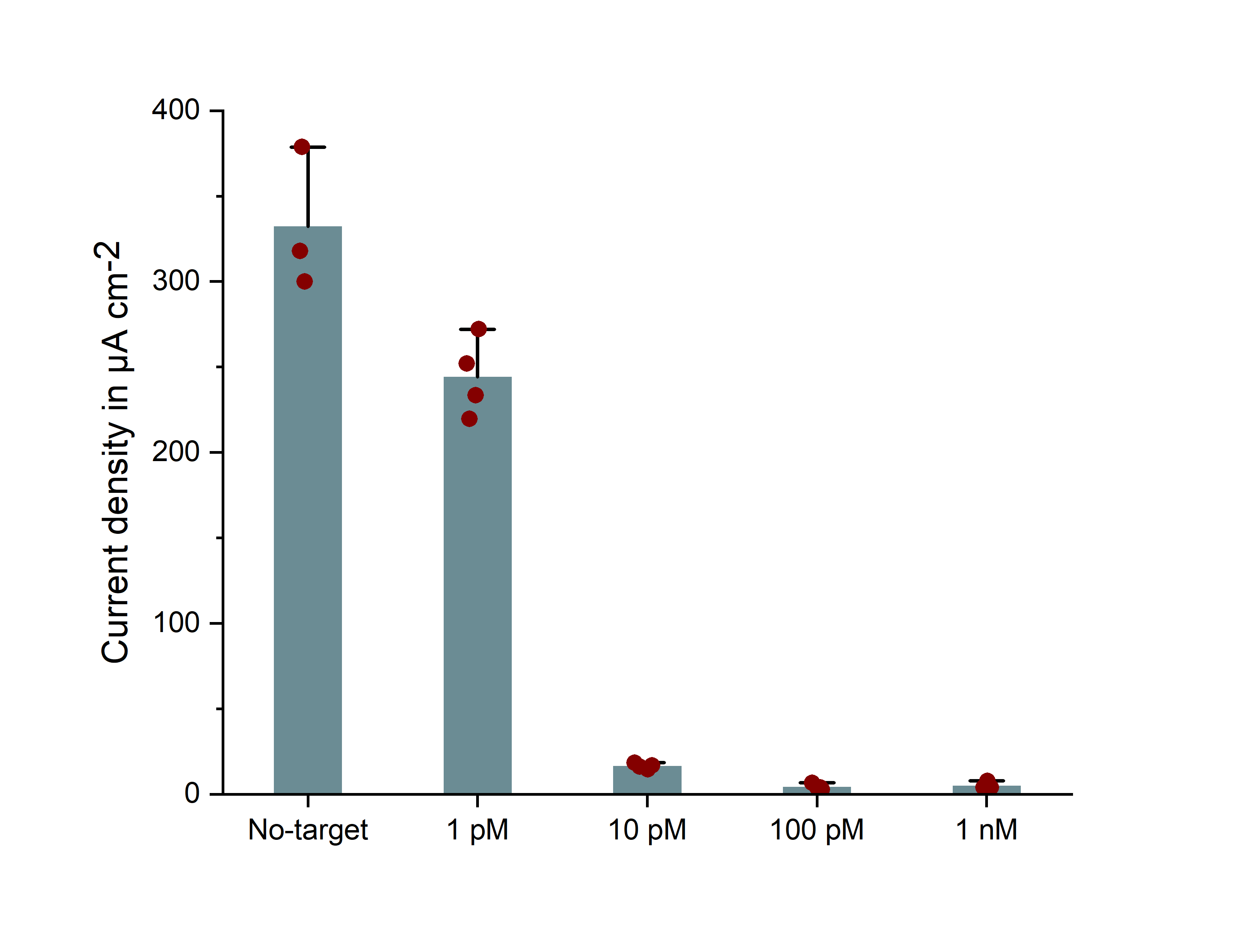

**Figure S3. Validation of the modified assay protocol as well as synthetic crRNA and target RNA for RdRP (SARS-SoV-2).** Proof-of-principle measurement of 4 different concentrations of the synthetic SARS-CoV-2 RdRP gene was successfully conducted using the standard experimental protocol (3-hour incubation of the sample solution at 37 °C; Error bars represent +SD of n = 4).

**Prolonged Cleavage Time**

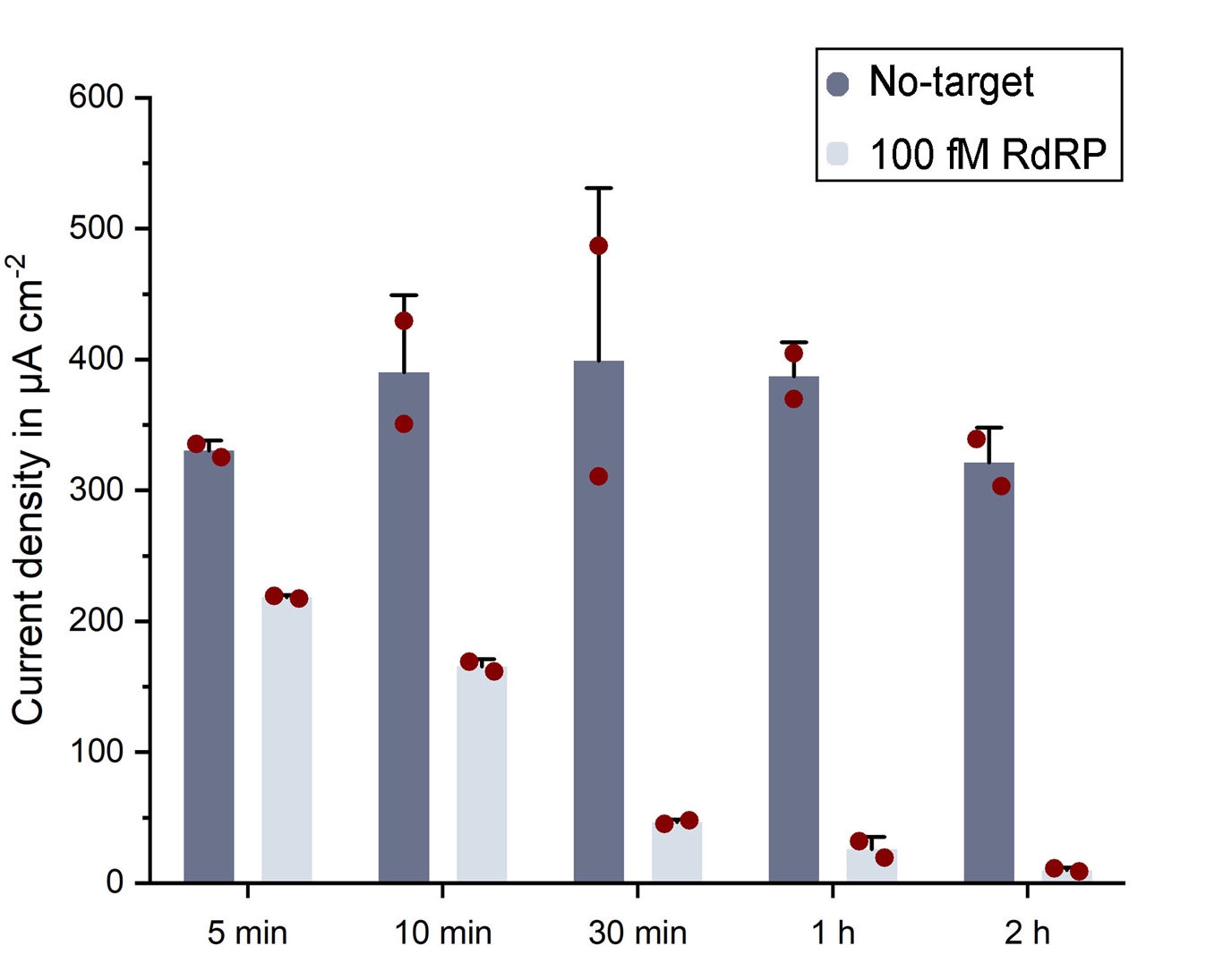

**Figure S4. Prolonging the incubation time of the sample solution can eliminate ambiguity in the amperometric readout.** When incubating the sample solution with 100 fM of the target sequence at 37 °C a sharp decrease in uncleaved reRNA and therefore in signal amplitude can already be observed within 25 minutes of additional (30 min) cleavage time. After 2 hours of all available reRNA has been cleaved and there is no more amperometric signal (Error bars represent +SD of n = 4).

**Glucose Oxidase Conjugation**

For the enzyme conjugation of the monoclonal anti-6-FAM IgG, CF™ 488A antibody (SAB4600050-125UL, Merck, Germany), we used the Glucose Oxidase (GOx) Conjugation Kit – Lightning-Link® (ab102887, Abcam, UK). In brief, the modifier reagent was added to the antibody unlabeled (1:11) which was then diluted (1:1) with PBS (10 mM, pH 7.4). Subsequently the mixture was added to the lyophilized GOx, resuspended by gentle pipetting and incubated at room temperature, in the dark for 3 hours. Afterwards, the quencher reagent (1:11 antibody) was added and the reaction vessel was left at room temperature (dark) for another 30 minutes. For a detailed description of the process, please refer to the manufacturers protocol booklet^1^.

**Oligonucleotides**

**Table S2. Summary of oligonucleotides and their specifications as used in this study.** Target site sequences were selected according to Corman et al.^2^.

| **Assay** | **Oligonucleotide** | **Sequence** | **Concentration** |
| --- | --- | --- | --- |
| E gene | Target | 5’–CGAAGCGCAGUAAGGAUGGCUAGUGU–3’ | x |
|  | crRNA | 5’–GGGGAUUUAGACUACCCCAAAAACGAAGGGGACUAAAACACACUAGCCAUCCUUACUGCGCUUCG–3’ | 62.5 nM |
| RdRP gene | Target | 5’–GCAUCUCCUGAUGAGGUUCCACCUG–3’ | x |
|  | crRNA | 5’–GGGGAUUUAGACUACCCCAAAAACGAAGGGGACUAAAACCAGGUGGAACCUCAUCAGGAGAUGC–3’ | 62.5 µM |
| RdRP gene  SARS-CoV-1 | Target | 5’–CCAAGGUGGAACAUCAUCCGGUGAUGC–3’ | x |
|  | crRNA | 5’–GGGGAUUUAGACUACCCCAAAAACGAAGGGGACUAAAACGCAUCACCGGAUGAUGUUCCACCUUGG–3’ | 62.5 µM |
| miR-520f | Target | 5‘–AAGUGCUUCCUUUUAGAGGGUU–3’ | x |
|  | crRNA | 5’–GGGGAUUUAGACUACCCCAAAAACGAAGGGGACUAAAACAACCCUCUAAAAGGAAGCACUU–3’ | 62.5 µM |
| miR-19b | Target | 5’–UGUGCAAAUCCAUGCAAAACUGA–3’ | x |
|  | crRNA | 5’–GGGGAUUUAGACUACCCCAAAAACGAAGGGGACUAAAACUCAGUUUUGCAUGGAUUUGCACA–3’ | 62.5 µM |
|  | reRNA_14b | 5'–5AUGGC55AUGGC5–3' | 250 nM |
|  | reRNA_20U | 5’–UUUUUUUUUUUUUUUUUUUU–3’ | 250 nM |

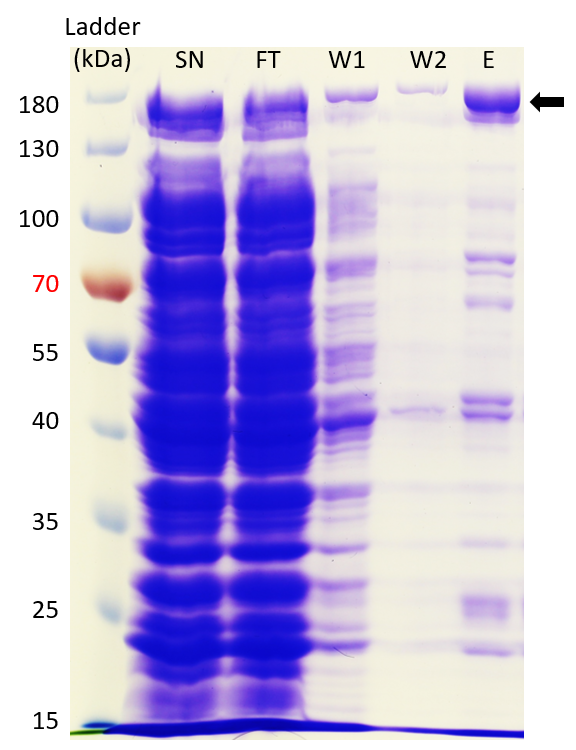
**SDS-PAGE**

**Figure S5. Analysis of LbuCas13a purification by SDS-PAGE.** 6 µl of PageRuler Prestained Protein Ladder (Thermo, lane 1) and 10 µl of samples were separated on a 12% (w/v) SDS-gel. P, pellet; SN, supernatant; FT, flow-through; W1, 1^st^ wash fraction; W2, 2^nd^ wash fraction; E, elution. The arrow indicates the LbuCas13a (expected size: 183.3 kDa).

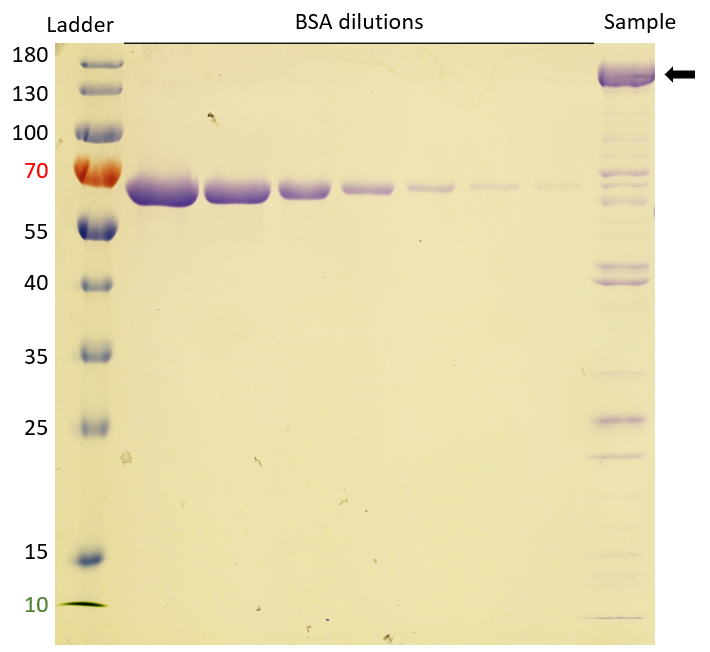
**Purity Determination**

**Figure S6. Purity determination of the LbuCas13a sample.** 6 µl of PageRuler Prestained Protein Ladder (Thermo, lane 1) and seven dilution series of bovine serum albumin (0.5 - 0.0078 mg ml^–1^), and the purified protein (1:2 diluted) were loaded on a 12% (w/v) SDS gel. From the pixel intensities the concentration was determined based on the calibration curve from the BSA dilutions.

**Cost Estimation of CRISPR-Biosensor**

**Table S3. Cost estimation for the biosensor based on the fabrication of four wafers.** Costs for assay components were based on one wafer, whereas the reagents for the MasterMix are calculated per reaction.

| Fabrication Step | Material / Reagent | miLab sensor  (130 sensors each) | 4/6-fold multiLab sensor  (30 / 26 sensors each) |
| --- | --- | --- | --- |
| Substrate | Pyralux® AP8545 | 0.097 € | 0.105 / 0.081 € |
| Lift-off process | ma-N 1420 | 0.024 € | 0.026 / 0.020 € |
| Platinum vapor deposition | Platinum | 0.480 € | 0.520 / 0.400 € |
| Electrode isolation | SU-8 3005 | 0.067 € | 0.073 / 0.056 € |
| Galvanic silver deposition | Ag/AgCl | < 0.001 € | < 0.001 € |
| Stopping barrier | 3% Teflon® | < 0.001 € | < 0.001 € |
| Dry-film photoresist | Pyralux® PC1025 | 0.01 € | 0.011 / 0.008 € |
|  |  | **0.680 €** | **0.737 / 0.567 €** |
| Assay Components | | |  |
| Surface functionalization | Neutravidin | 0.074 € | 0.148 / 0.222 € |
| Surface blocking | Casein | 0.008 € | 0.016 / 0.024 € |
| Blocking of NA | Biotin | 0.364 € | 0.728 / 1.092 € |
| Enzyme labelling | Ab-GOx | 0.031 € | 0.062 / 0.093 € |
|  |  | **0.467 €** | **0.954 / 1.431 €** |
| Total | | **1.147 €** | **1.691 / 1.998 €** |
| MasterMix |  | **Cost per reaction** | |
| reRNA_20U | 6-FAM and biotin labeled RNA | 0.018 € | |
| crRNA | Custom oligonucleotide | < 0.001 € | |
| Inhibitor | Murine RNase Inhibitor | 0.072 € | |
| Effector | LbuCas13a | 0.063 € | |
| Total | | **0.154 €** | |

**Multiplexed Chip Design and Measurements**

**a**

**b**

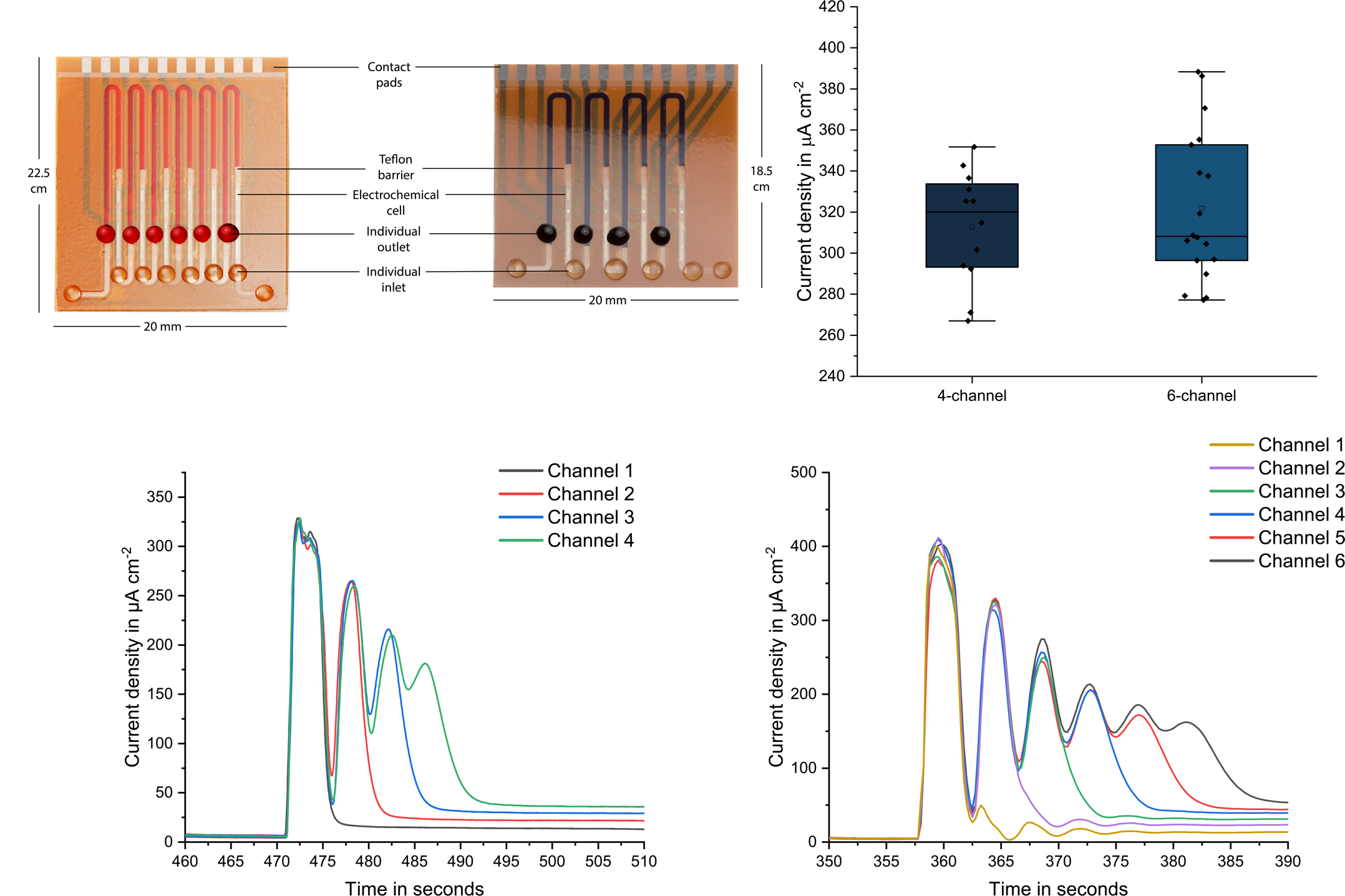

**c**

**d**

**Figure S7. a) Photos of the implemented single-channel multiplexed biosensors**. Each one contains either four or six consecutive incubation areas, individual electrochemical cells and Teflon barriers preventing electrode fouling. **b)** Performance test conducted with a model assay, where channels are incubated with 200 µg ml^–1^ Streptavidin-GOx for 20 minutes. **c-d)** **Exemplary amperometric signal readout of the multiplexed biosensor** with 4 (c) and 6 (d) incubation areas. The first four successive peaks correspond to the amount of electrochemically active species that have accumulated within the immobilization areas during the stop-flow protocol. In the final “flow” phase of the protocol, these species are also passing through their subsequent neighboring electrochemical cells in addition to their own individual ones, which creates the following peaks.

**a**

**b**

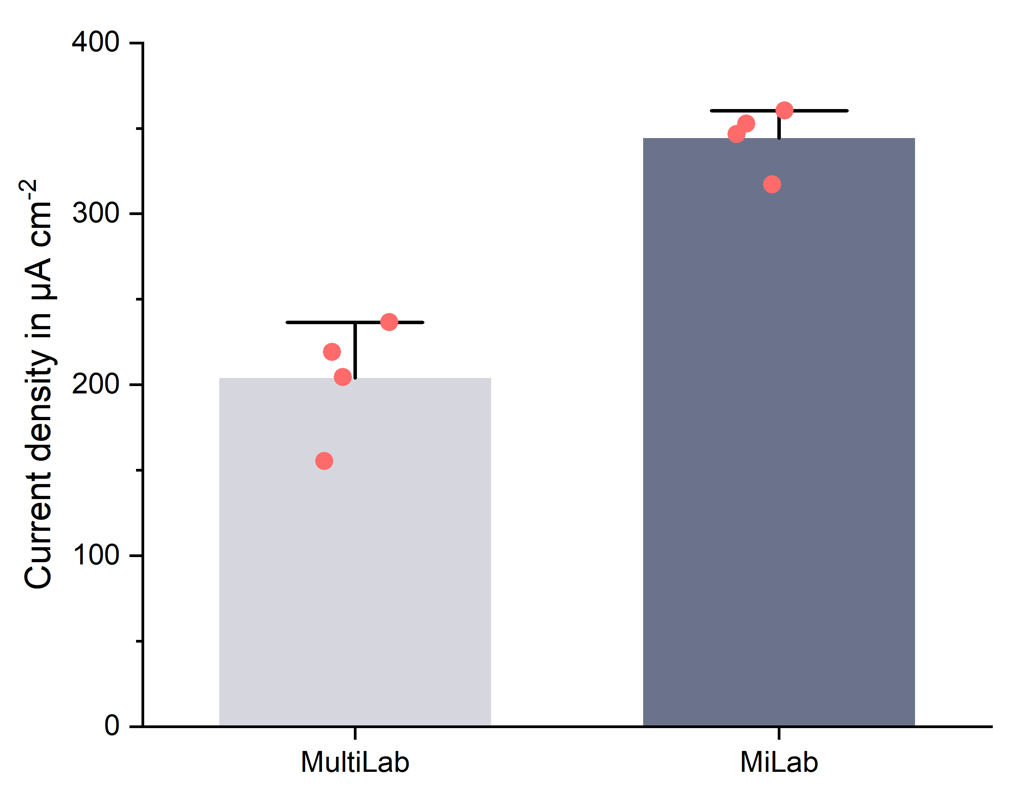

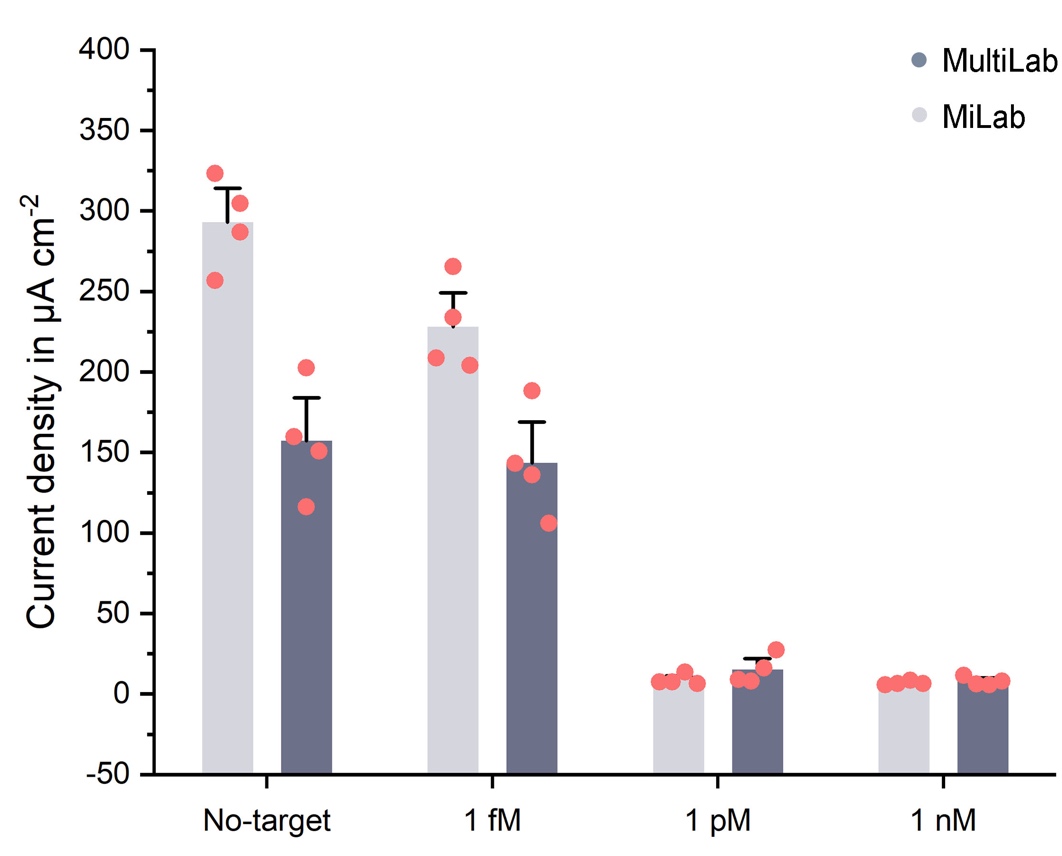

**Figure S8. a) Calibration of the CRISPR/Cas13a powered assay on a 4-analyte multiplexed biosensor.** Measurement of 3 dilutions of the SARS-CoV-2 E gene for comparison between the single-analyte and multiplexed platforms. **b) Signal comparison of the model assay on 2 miLab chips *versus* the multiLab device.** The observed signal difference for hydrogen peroxide oxidation between both platforms is caused by the different flow rates applied (milab at 20 µl min^–1^ & multiLab at 10 µl min^–1^) (Bar plot for n = 4 replicates (red dots signify sensors), error bars represent ± SD).

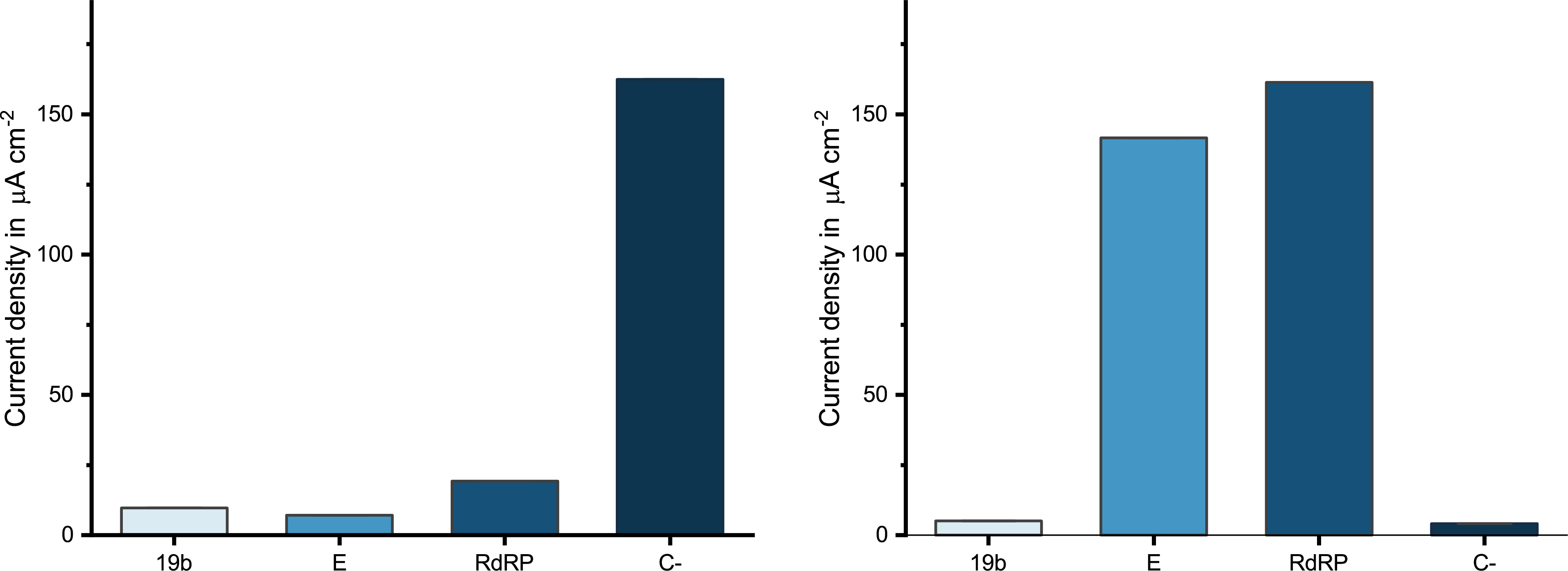

**b**

**a**

**Figure S9. Proof-of-principle for the ability fo discriminate between COVID-19 and SARS using a multiplexed biosensor. a)** Mock patient 1: infected with SARS-CoV-2 and **b)** mock patient 2: infected with SARS-CoV-1.

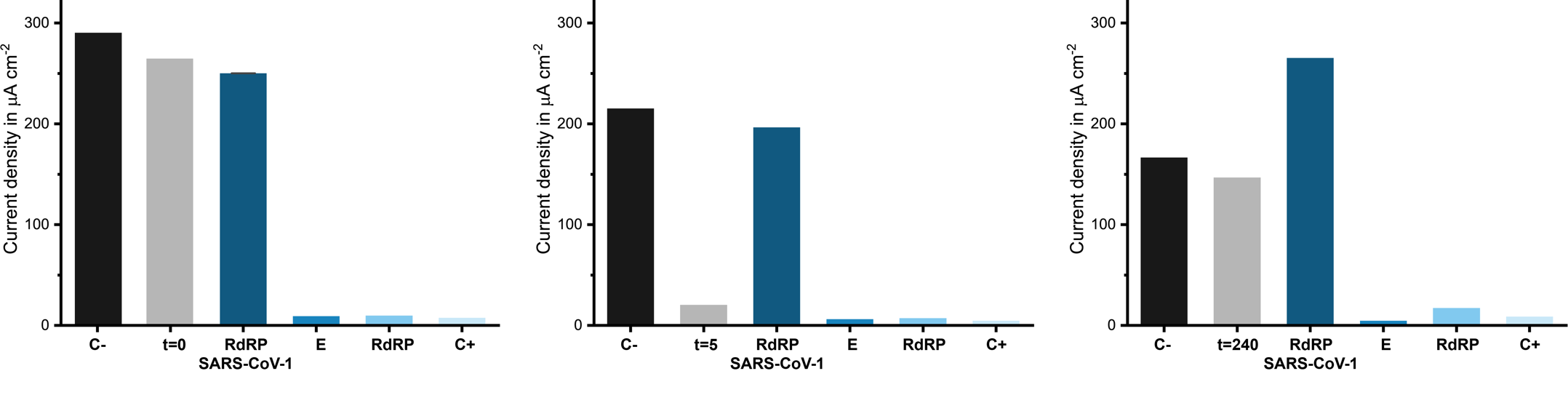
**Figure S10.** **Personalized antibiotic dosing in a mock COVID-19 patient.** Measured current densities **a)** before, **b)** 5 minutes and **c)** 240 minutes after administration of the antibiotic.

**b**

**c**

**a**

**Further Miniaturization of the Measurement Setup**

**b**

**c**

**a**

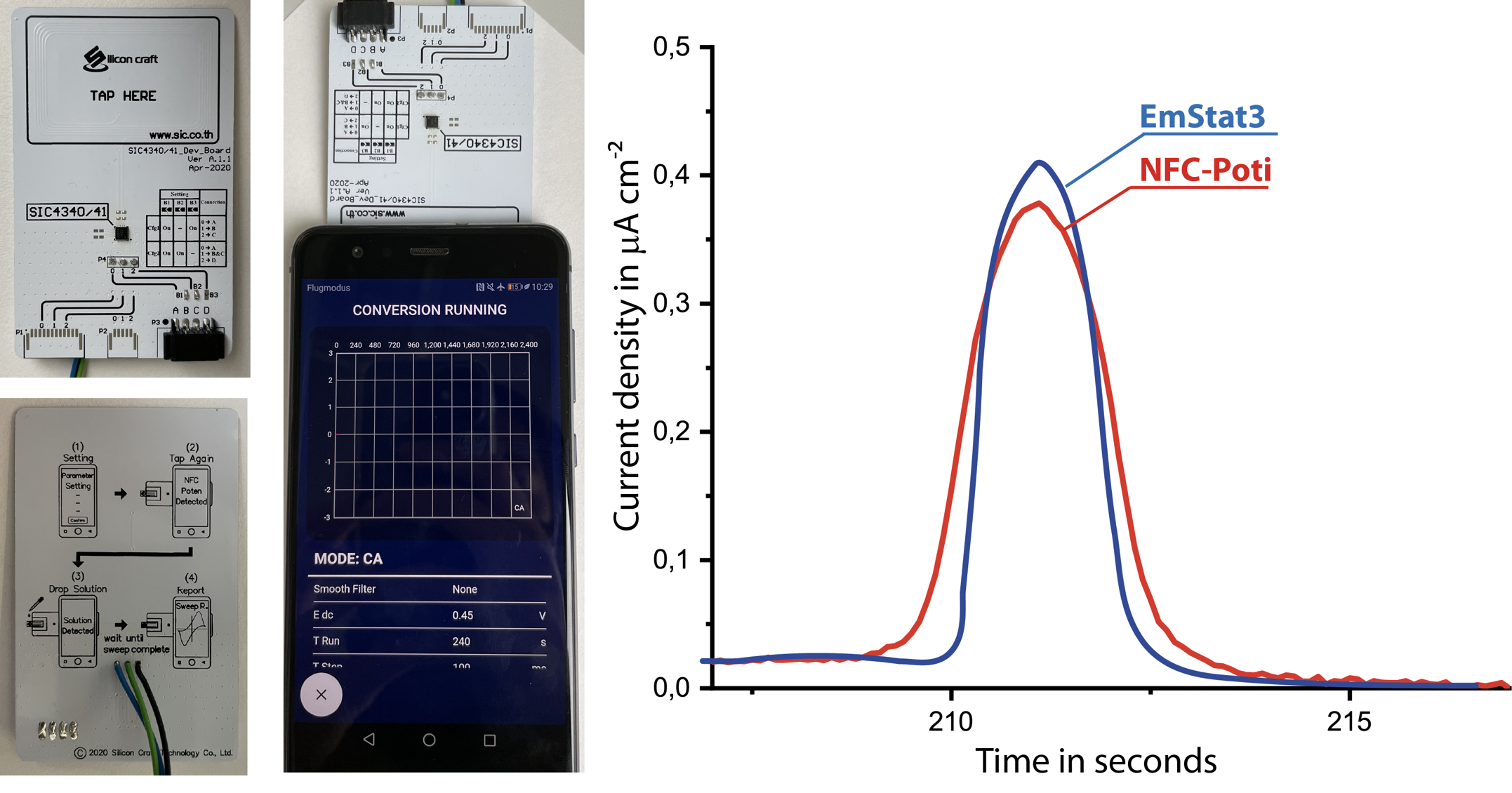

**Figure S11. Demonstration of the point-of-care applicability of the CRISPR-Biosensor platform using an NFC potentiostat.** **a)** NFC potentiostat card from front and backside. **b)** Smartphone connection and the measurement software. **c)** Comparison of the performance of the NFC potentiostat (SIC4341 – Potentiometric sensor interface with NFC type 2, Silicon Craft Technology PLC, Thailand) with a commercial bench-top potentiostat (EmStat3; PalmSens, The Netherlands).

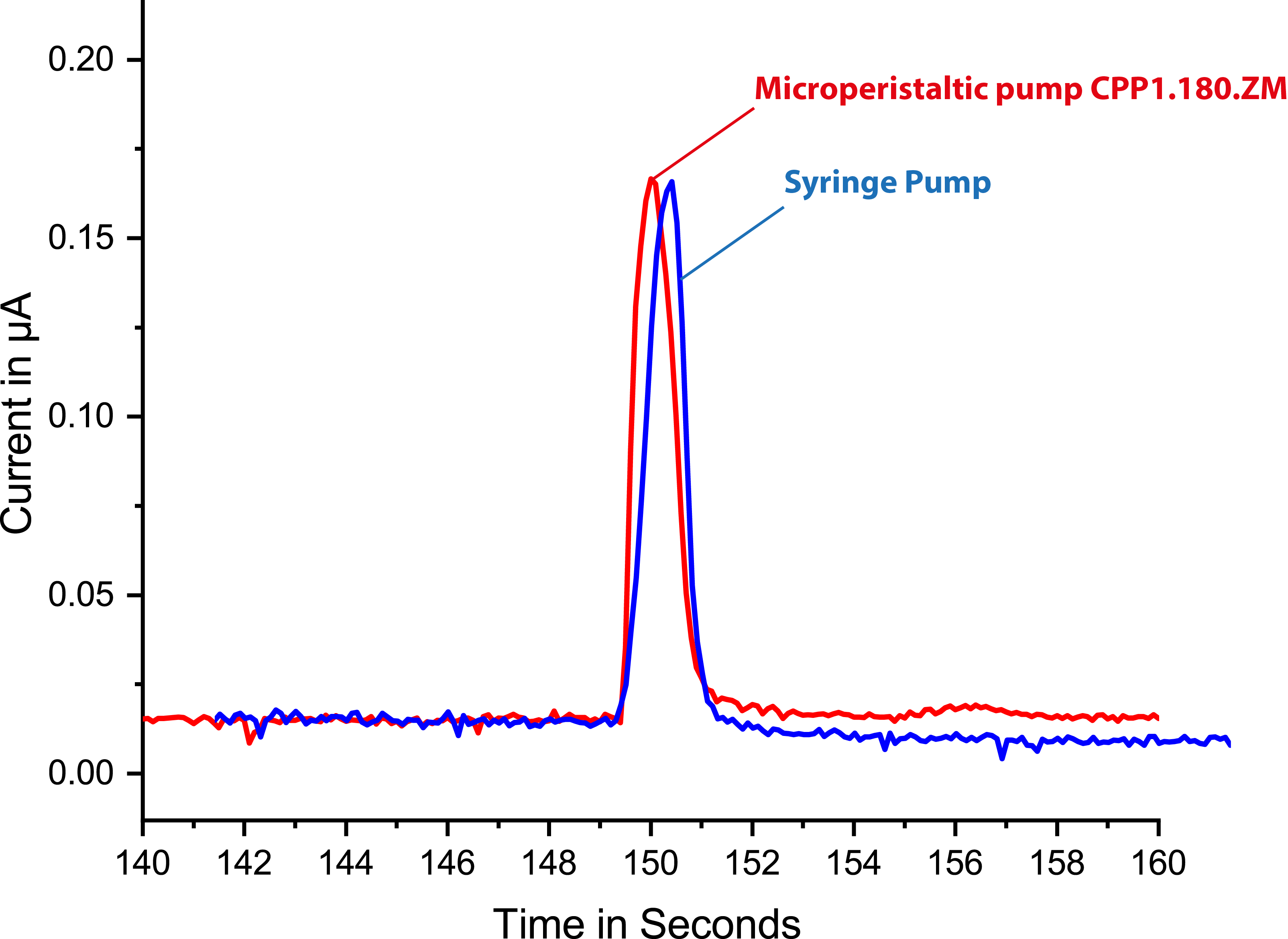

**Figure S12. Performance comparison with respect to fluidic control between a microperistaltic pump (**[**MP.CPP1.180.ZM**](http://mp.cpp1.180.zm/)**, Jobst Technologies, Germany) and a syringe pump (PHD ULTRA™ 4400, Harvard Apparatus, USA).** The measurement was performed using single-channel biosensor incubated with model assay (100 µg ml^–1^ Avidin-GOx). Signal readout is achieved using NFC potentiostat (SIC4341 – Potentiometric sensor interface with NFC type 2, Silicon Craft Technology PLC, Thailand) by applying 2-min stop-flow protocol.

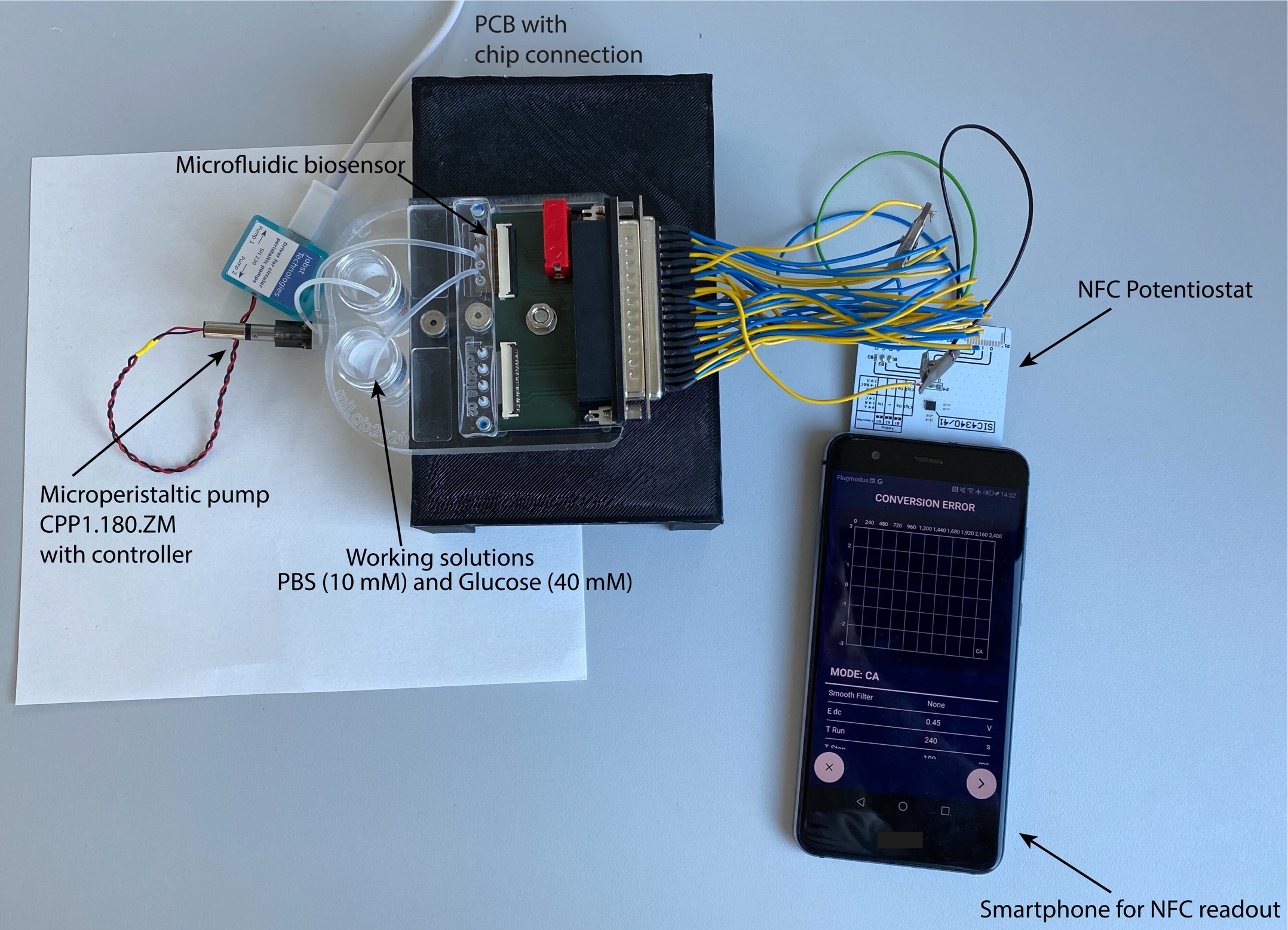

**Figure S13. Demonstration of the point-of-care applicability of the CRISPR-Biosensor platform.** Measurement setup includes a microperistaltic pump, a custom-made chip holder, an NFC-potentiostat, the microfluidic biosensor and a signal readout unit (i.e., smartphone).
