## Supplementary material for "Multiplexed biosensor for point-of-care COVID-19 monitoring: CRISPR-powered unamplified RNA diagnostics and protein-based therapeutic drug management": Statement of ethic commission

Ethik-Kommission ◦ Engelberger Straße 21◦ 79106 Freiburg

Albert-Ludwigs-Universität Freiburg
Department of Microsystems Engineering (IMTEK)
Dr.-Ing. Can Dincer
Georges-Köhler-Allee 103
79110 Freiburg

Albert-Ludwigs-Universität Freiburg

**Ethik-Kommission**

Prof. Dr. R. Korinthenberg
(Vorsitzender)
M. Schmidt
(Geschäftsführer)

Engelberger Straße 21
79106 Freiburg

Tel. 0761/270-72500
Fax 0761/270-72630

www.ethik-kommission.uniklinik-freiburg.de


Bearbeitet von:
Jonas Christoph

**Freiburg,** 29.03.2022

| | |
|---|---|
| **Application Number:** | 22-1137-Anfrage **(***please quote in any communication)* |
| **Project Leader:** | Dr.-Ing. Can Dincer |
| **Research Project:** | Methodenvalidierung: Analyse von COVID-19-Patientenproben<br>[Method validation: analysis of COVID-19 patient samples]<br>Your request dated 14th March 2022 |

Dear Dr. Dincer,

thank you for your request dated 14th March 2022 regarding the research project mentioned above.

The Ethics Committee hereby declares that there was no obligation for you to seek advice from the Ethics Committee under German law.

Yours sincerely

R. Korinthenberg

Prof. Dr. R. Korinthenberg
Chairman of the Ethics Committee
